# Learning the shared structure of human health across diseases, modalities, and time

**DOI:** 10.64898/2026.07.07.26357373

**Authors:** Paul Hager, Benedikt Roth, Niklas Bühler, Diyuan Lu, Jamison H. Burks, Liubov Shilova, Raphael Kfuri-Rubens, Eljas Roellin, Jiazhen Pan, Maxime Di Folco, Emily Chan, Julia A. Schnabel, Lisa Adams, Daniel Rueckert, Fabian J. Theis, Francesco Paolo Casale

## Abstract

Human disease risk emerges from the shared influences of genetics, environment, lifestyle, and concurrent diseases over time, resulting in recurring patterns of susceptibility across conditions. However, most risk prediction models treat diseases as independent outcomes or rely on limited input variables, restricting their ability to capture these shared patterns. Here we present RisQ, a framework that learns a unified representation of human health across diseases, modalities, and time. This representation is queried with natural language to estimate disease risk for arbitrary diseases and prediction horizons. Generalization to unseen disease groups and prediction horizons indicates that information is shared across diseases and time, revealing a common structure of disease risk that is learnable. Trained and validated in 488,170 participants from the UK Biobank and evaluated without retraining in 257,538 participants from the independent All of Us cohort, RisQ leverages this shared structure to outperform disease-specific models, multi-disease frameworks, and tabular foundation models in risk prediction. We show that jointly modeling increasing numbers of diseases, input modalities, and prediction horizons improves performance, indicating that scaling these axes increases information transfer and enriches the learned structure. We then show this structure is multi-scale: it captures demographic determinants of disease susceptibility, while also organizing individuals into reproducible cross-disease risk clusters within demographically restricted subgroups. Genetic analyses further support the biological grounding of the structure by linking gene-level loss of function to cross-disease risk profiles. This surfaces known relationships of *HBB*, *SLC22A12*, *CASR*, and *LDLR*, while also highlighting less characterized associations. Together, these results indicate that human disease risk exhibits a shared structure that can be learned from multimodal data to improve risk prediction, stratify individuals by cross-disease susceptibility, and support the discovery of relationships across diseases.

## 1 Introduction

The human body is an interconnected system in which physiological subsystems do not operate independently ^1–3^. As a result, diseases frequently co-occur and influence one another, even when they appear unrelated ^4–7^. At the same time, disease risk is shaped by a wide range of factors spanning genetics, environment, and lifestyle, and evolves over time with age ^8–13^. Together, these observations suggest that an accurate representation of health should capture the joint structure linking diseases, risk factors, and time.

Despite strong biological evidence, many predictive models of disease have not kept pace. Most prior work focuses on limited disease endpoints and time horizons, even when leveraging diverse risk factors ^14–16^. While effective in isolation, this narrow scope limits the ability to capture shared signal and benefit from scaling data and model size ^17^. A complementary line of work shows that jointly modeling many diseases can improve prediction ^18^, but relies primarily on diagnostic history, leaving substantial signal in biomarkers, environment, genetics, and other modalities untapped. What remains lacking is a framework that learns from multimodal data, predicts across diseases, and estimates risk continuously over time, leveraging shared patterns to improve personalized prediction and enable biological insight.

To this end, we introduce RisQ, a transformer-based multimodal framework that learns a unified representation of human health across diseases, modalities, and time. We train and validate RisQ on 488,170 individuals from the UK Biobank (UKB) ^19^, integrating diagnostic history, medications, biomarkers, physical measurements, lifestyle factors, environmental exposures, and genetics to predict risk for over 1,000 diseases across 16 years of follow-up.

RisQ generalizes to unseen disease chapters and extrapolates to unseen time horizons through its query mechanism, which combines natural language disease targets with continuous representations of time. This indicates that information is shared across diseases and time horizons, revealing a common structure of disease risk that can be learned. By leveraging this shared structure, RisQ outperforms existing models in both the UKB and the independent All of Us cohort, where we evaluate generalization without retraining under reduced input availability. In extensive analyses, we show that increasing disease targets, temporal supervision, and multimodal inputs each independently improves predictive performance, indicating that scaling these axes enables better information transfer and improved structure. We show that this learned structure captures both broad demographic determinants of disease susceptibility as well as reproducible cross-disease risk profiles within demographically coherent subpopulations, and links gene-level loss-of-function variation to established cross-disease risk profiles.

## 2 Results

### 2.1 Learning a shared representation of disease risk

RisQ is a transformer-based framework that encodes multimodal participant data into a unified representation of health. This representation supports prediction of disease risk across conditions and time, enabling a single model to estimate risk for any disease over arbitrary horizons (**Fig. 1a**).

**Figure 1.**
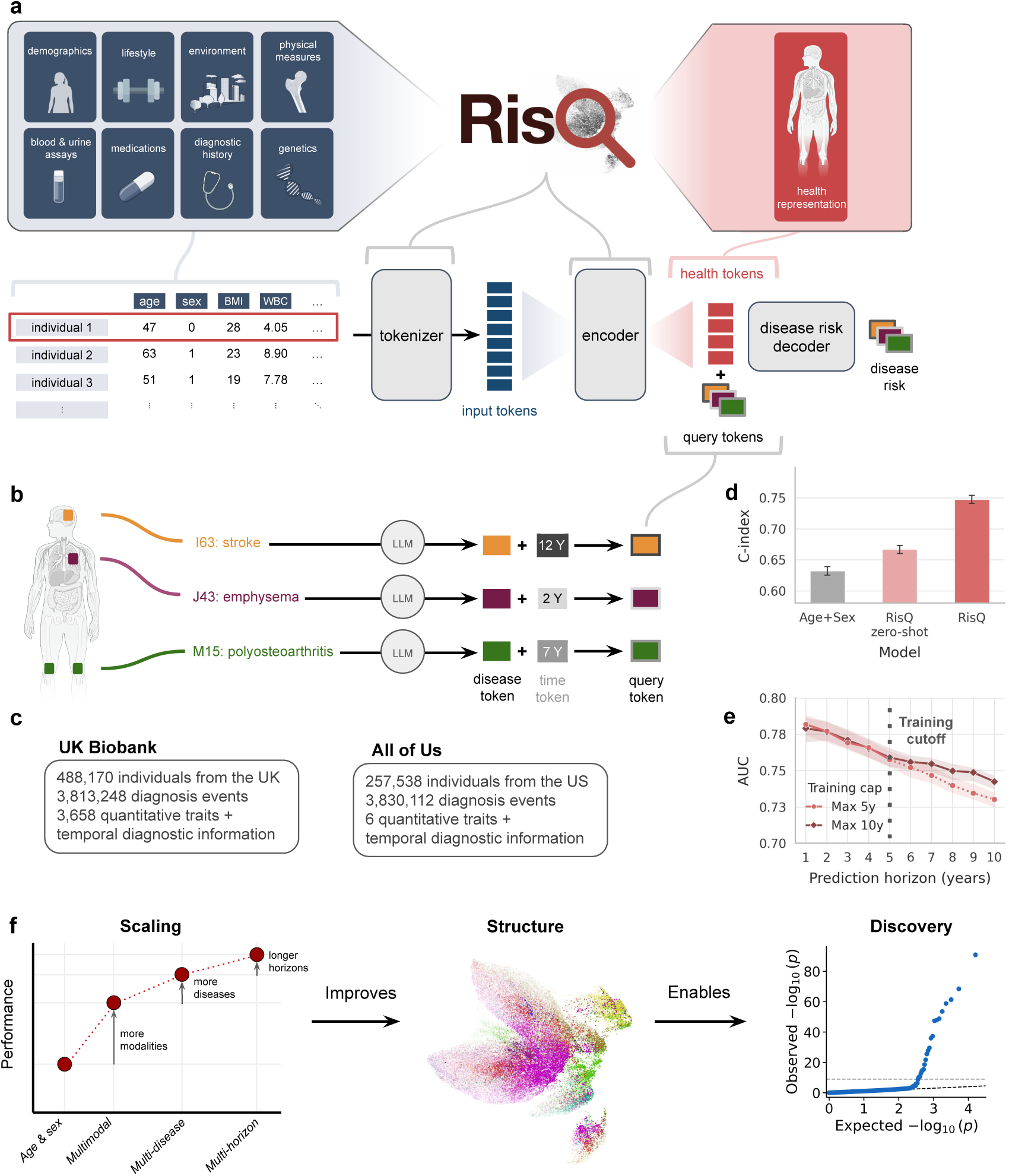
| RisQ learns a structured representation of health for multi-disease risk prediction and biological discovery. (**a**) Multimodal data including demographics, lifestyle, environmental exposures, physical measurements, blood and urine assays, medications, diagnostic history, and genetics, are tokenized and encoded into a condensed set of health tokens. Query tokens, which are conditioned on disease and time, interact with the health tokens through a disease risk decoder to generate disease-risk predictions. (**b**) Disease-risk queries specify both the disease of interest and the prediction horizon. (**c**) RisQ was trained and validated on 488,170 participants from the UK Biobank and evaluated without retraining on 257,538 participants from the independent All of Us cohort. (**d**) RisQ generalizes to unseen disease chapters: macro C-index on 569 disease targets withheld from RisQ zero-shot training in turn under a leave-one-chapter-out protocol, comparing RisQ zero-shot, fully supervised RisQ, and fully supervised RisQ restricted to age and sex inputs. Error bars denote 95% bootstrap confidence intervals. (**e**) RisQ extrapolates beyond its training horizon: a model trained with a five-year observation cap (light red) is evaluated at horizons up to ten years and compared to a ten-year-capped reference (dark red). Solid lines indicate horizons within the training cap; dotted lines indicate extrapolation. Shaded bands indicate 95% confidence intervals. (**f**) Scaling modalities, diseases, and time horizons improves the disease risk structure learned by RisQ. The structure can be used for risk stratification and enables biological discovery. Unless otherwise noted, error bars and shaded bands indicate 95% confidence intervals (2.5th–97.5th percentiles; *n* = 100 resamples).

RisQ integrates structured variables, medications, and longitudinal diagnostic histories. Structured inputs are embedded using feature-specific tokenizers ^20^, while diagnoses and medications are encoded with large language models ^21^, with temporal information incorporated via continuous sinusoidal encodings (**Methods**). These tokens are encoded into a fixed set of health tokens (**Fig. 1a**), yielding a unified representation.

This unified representation can be queried to estimate disease risk for specific diseases and time horizons. Each query is represented by a token constructed by combining an LLM embedding of the disease with a temporal token specifying the prediction horizon (**Fig. 1b**; **Methods**). The risk estimate is then extracted by a disease risk decoder that takes as input both the health tokens and the query token. By training to predict risk across queries spanning diseases and prediction horizons, RisQ learns representations that enable information transfer across endpoints and timescales within a unified model, allowing estimation of both short- and long-term risk (e.g., two-year risk of acute myocardial infarction and fifteen-year risk of Alzheimer’s disease) from a single representation.

We trained and validated RisQ on 488,170 UKB participants, of which 33,705 were held-out test subjects, integrating 3.8M diagnosis events with 3,658 structured and unstructured features spanning medications, biomarkers, physical measurements, environmental exposures, lifestyle variables, and genetics (**Fig. 1c**; **Methods**), finding RisQ to scale well with increasing data (**Supplementary Fig. S1a**). We further assess generalization under cohort and input shift through external validation without retraining in 257,538 participants from the independent All of Us cohort (**Fig. 1c**), described in the following section.

We evaluated whether predictive information is shared across diseases and time by testing generalization to unseen disease groups and prediction horizons through the query mechanism. We withheld individual ICD chapters during training and evaluated disease risk for these unseen targets. RisQ achieved strong zero-shot out-of-chapter performance (C-index = 0.667, 95% CI 0.660–0.673), substantially outperforming a demographic baseline using age and sex alone (C-index = 0.632, 95% CI 0.625–0.639, *P* = 1.40 *×* 10*^−^*^8^; **Fig. 1d**; **Supplementary Fig. S2d**), indicating that predictive signal transfers across disease groups. We next evaluated temporal extrapolation by restricting observed prediction horizons during training and assessing performance at longer horizons (**Fig. 1e**; **Methods**). A model trained with supervision limited to five years achieved a macro AUC of 0.730 (95% CI 0.725–0.735) at an unseen ten-year horizon, retaining 95% of the above-chance performance of a reference model trained with ten-year supervision (macro AUC = 0.742, 95% CI 0.737–0.747).

Together, these results show that information is shared across diseases and time horizons, supporting a common structure of disease risk that can be learned. We next show that leveraging this shared structure improves risk prediction across diseases, and that prediction gains increase when scaling diseases, modalities, and prediction horizons, consistent with greater information transfer across these axes and richer disease-risk structure. We then characterize this structure and show its usefulness for risk stratification and biological discovery (**Fig. 1f**).

### 2.2 Shared representations improve disease-risk prediction

We next evaluated whether leveraging this shared structure in disease risk improves risk prediction relative to existing multi-disease and disease-specific models. RisQ was assessed in a held-out UKB cohort of 33,705 participants across 588 clinician-curated diseases, excluding congenital, pregnancy-related, perinatal, injury, and external-cause codes (**Supplementary Dataset 1**; results for all 1,246 diseases are shown in **Supplementary Fig. S2**). Models were evaluated on binary classification of disease occurrence at 1-, 2-, 5-, and 10-year prediction horizons and survival analysis until censoring (maximum 16 years; **Methods**). We compared RisQ against the multi-disease forecasting model Delphi-2M ^18^, the tabular foundation model TabPFNv3^22^, and disease-specific XGBoost ^23^ models under both default and extensively optimized configurations.

Across all horizons, RisQ achieved the highest discrimination (1-year AUC 0.791, 95% CI 0.783–0.800; 2-year 0.790, 0.784–0.796; 5-year 0.768, 0.761–0.775; 10-year 0.760, 0.753–0.766; Bonferroni-corrected *P <* 3.3 *×* 10*^−^*^3^ across horizons and baselines; Fig. 2a; **Supplementary Fig. S3**) and the highest C-index in survival evaluation (0.760, 95% CI 0.754–0.766, *P* = 0.043 across matched diseases vs. the optimized XGBoost Survival; Fig. 2b), while being well-calibrated (ECE *≤* 0.002; Brier score *≤* 0.006; **Supplementary Fig. S4**). Improvements were observed relative to both multi-disease and disease-specific baselines, including optimized XGBoost models despite their greater computational costs (**Supplementary Fig. S1b**). RisQ ranked highest by median AUC in the majority of ICD-10 chapters across all horizons (11/12 and 10/12 chapters at the 2- and 10-year horizons, respectively; Fig. 2c; **Supplementary Fig. S3a**), indicating that performance is consistent across clinically distinct disease categories and time horizons.

**Figure 2.**
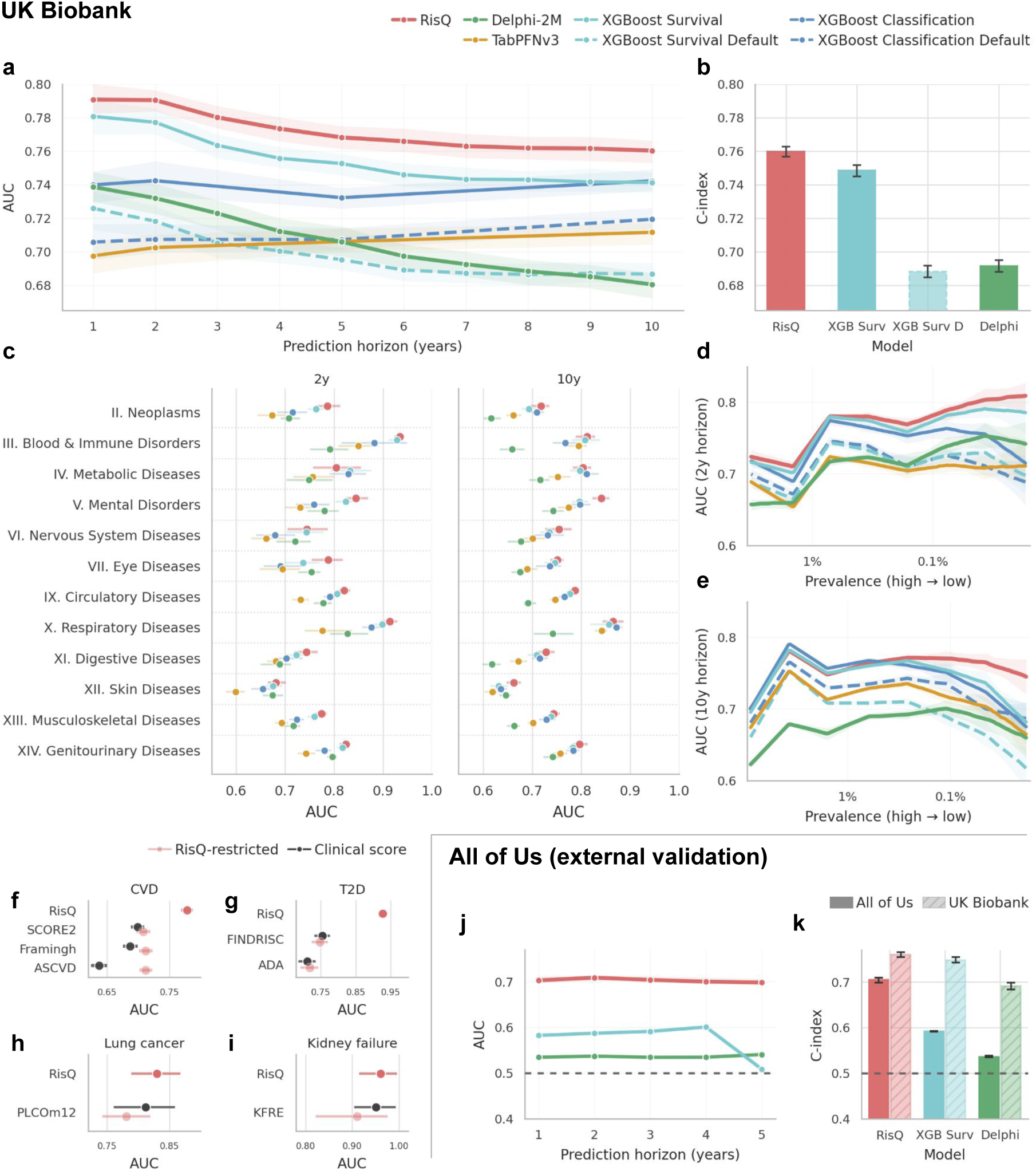
| Shared multimodal representations improve disease-risk prediction. (**a**) Macro AUC across prediction horizons (1–10 years) in the held-out UK Biobank test split; RisQ outperforms foundation-model and gradient-boosted baselines at all horizons. (**b**) Survival evaluation (C-index) shows superior discrimination of RisQ relative to survival baselines. (**c**) Macro AUC across ICD-10 chapters at the 2- and 10-year horizons demonstrate consistent performance gains across clinically distinct disease categories. (**d,e**) AUC stratified by disease prevalence at 2-year (**d**) and 10-year (**e**) horizons; prevalence is shown on a log scale. (**f–i**) Comparison with established clinical risk scores for cardiovascular disease (**f**), type 2 diabetes (**g**), lung cancer (**h**), and kidney failure (**i**); “RisQ-restricted” denotes evaluation using the same input variables as each clinical score. (**j,k**) External validation in the All of Us cohort under zero-shot evaluation and reduced inputs showing classification performance across horizons (**j**) and survival performance (**k**). C-indices were computed independently within each cohort over the available follow-up (UKB: up to 16 years; All of Us: up to 5 years), and diseases were matched across models within each cohort separately. Unless otherwise noted, error bars and shaded bands indicate 95% confidence intervals (2.5th–97.5th percentiles; *n* = 100 resamples).

The relative advantage of RisQ increased as disease prevalence decreased (Fig. 2d**,e**). For low-prevalence diseases (< 0.1% event rate in the test set), performance gains over disease-specific models were most pronounced (2-year horizon: RisQ AUC= 0.800 vs. XGBoost-Surv AUC= 0.770, *P* = 4.37 *×* 10*^−^*^5^; 10-year horizon: RisQ AUC= 0.754 vs. XGBoost-Surv AUC= 0.708, *P* = 8.07 *×* 10*^−^*^9^; **Supplementary Fig. S5a,b**), consistent with improved predictions in low-data regimes where information sharing helps most.

To evaluate the coherence of predictions, we examined whether they are informed by clinically meaningful signals using Integrated Gradients ^24^. Established risk factors emerged as dominant contributors for exemplar diseases, including smoking for lung cancer, HbA1c for type 2 diabetes, and vascular and age-related features for heart failure (**Supplementary Fig. S6**). Across outcomes, recurrent contributors included age, prior diagnoses, and lifestyle variables, while disease-specific features were enriched for their corresponding endpoints.

We next compared RisQ to established disease-specific clinical risk scores, including three cardiovascular disease scores (Framingham ^25^, ASCVD ^26^, SCORE2-type ^27^), two type 2 diabetes scores (FINDRISC ^28^, ADA-type ^29^), one lung cancer model (PLCOm2012^30^), and the Kidney Failure Risk Equation (KFRE) ^31^. The full RisQ model outperformed all evaluated scores, with the largest gains observed in cardiovascular disease and diabetes prediction (Fig. 2f**–i**). When restricted to the inputs used by each clinical score (RisQ-restricted), performance matched that of the corresponding scores for these endpoints, with greater uncertainty for lung cancer and kidney failure due to limited case counts (Fig. 2f**–i**).

### CVD: cardiovascular disease, T2D: type 2 diabetes, Framingh: Framingham

Finally, we assessed generalization beyond the UKB dataset by evaluating zero-shot prediction in the All of Us cohort, a United States–based study, using a reduced feature set comprising diagnosis history together with age, sex, BMI, and smoking and drinking behaviors (**Methods**). Despite differences in participant composition, baseline disease frequency, recruitment structure, and input variables ^32^, RisQ maintained discrimination across 1–5 year horizons, with AUC *≥* 0.69 at all evaluated time points (1-year AUC 0.703, 95% CI 0.698–0.712; 2-year 0.709, 0.703–0.714; 5-year 0.698, 0.698–0.707; Fig. 2j), outperforming both XGBoost Survival (1-year AUC 0.582, 95% CI 0.580–0.584; 2-year 0.587, 0.586–0.588; 5-year 0.509, 0.502–0.516) and Delphi-2M (1-year AUC 0.535, 95% CI 0.532–0.537; 2-year AUC 0.537, 0.535–0.539; 5-year 0.541, 0.539–0.542) significantly (Bonferroni-corrected *P <* 10*^−^*^43^ across horizons and baselines). RisQ exhibited the smallest decline in macro C-index from UKB to All of Us, decreasing by 0.055 points versus 0.155 for Delphi-2M and 0.157 for XGBoost Survival (macro C-index computed over the maximally matched disease set within each cohort independently: 486 diseases on UKB, 280 on All of Us; Fig. 2k).

Together, these results show that leveraging shared structure across diseases, modalities, and time improves disease-risk prediction. RisQ achieves strong performance across diverse diseases and prediction horizons, while generalizing to external datasets under substantial distribution shift and input constraints.

### 2.3 Scaling across diseases, time, and modalities increases information transfer and improves risk prediction

We next asked whether information transfer scales with increasing diseases, time, and input modalities. This should improve prediction performance and would imply richer disease-risk structure. To test this, we compared the fully integrated model to variants restricted along each axis.

We first evaluated whether joint modeling across diseases is required for optimal performance by comparing the fully integrated multi-disease model to disease-specific models trained independently for each outcome. Joint training improved survival discrimination (multi-disease macro C-index 0.757, 95% CI 0.743–0.767 vs 0.634, 0.621–0.668 for single-disease; multi-disease C-index exceeded single-disease training on 484 out of 545 diseases with at least one positive test case, *P* = 5.25 *×* 10*^−^*^69^; paired Wilcoxon signed-rank test; Fig. 3a; **Supplementary Fig. S5c,d**), indicating that jointly modeling diseases allows for cross-disease learning.

The benefits of cross-disease modeling should be greatest for rare diseases, where individual endpoints provide limited training signal. To evaluate this, diseases in the held-out test set were ranked by prevalence and assessed cumulatively from the rarest 20% to the full panel. The multi-disease model outperformed single-disease models across all five prevalence-ranked subsets (BH-FDR *<* 0.05), with the largest margin among the rarest diseases (macro C-index 0.753, 95% CI 0.707–0.814 vs 0.518, 0.485–0.563, *P* = 8.59 *×* 10*^−^*^8^ for the rarest 20%; paired Wilcoxon signed-rank test; Fig. 3b). Moreover, progressively increasing the number of diseases included during training yielded monotonic improvements on the same rare-endpoint panel (**Supplementary Fig. S7a**), supporting cumulative benefits of cross-disease modeling.

We next examined whether shared temporal structure contributed to RisQ’s performance gains. To do so, we trained models using only 2, 5, or 10 years of follow-up and compared them with the full model trained across all 16 years. Extending the duration of outcome supervision progressively improved discrimination (Fig. 3c; macro C-index = 0.717, 95% CI 0.710–0.723; 0.737, 0.732–0.743; 0.747, 0.740–0.753; and 0.749, 0.744–0.755 for training up to 2, 5, 10, and 16 years, respectively). Uncapped training (16-year supervision) yielded higher per-disease C-index than each capped variant (*P* = 2.37 *×* 10*^−^*^46^, *P* = 6.03 *×* 10*^−^*^16^, and *P* = 0.010 vs 2-, 5-, and 10-year caps, respectively; BH-FDR *<* 0.05 across the three comparisons; paired Wilcoxon signed-rank test). Evaluation across specific prediction windows further showed the benefits of multi-horizon training. Models trained with shorter follow-up retained meaningful discrimination beyond the time span observed during training, whereas longer supervision improved performance even at shorter horizons (**Supplementary Fig. S7b**). This suggests that disease-risk structure is shared across timescales, allowing information learned at one horizon to improve prediction at others.

**Figure 3.**
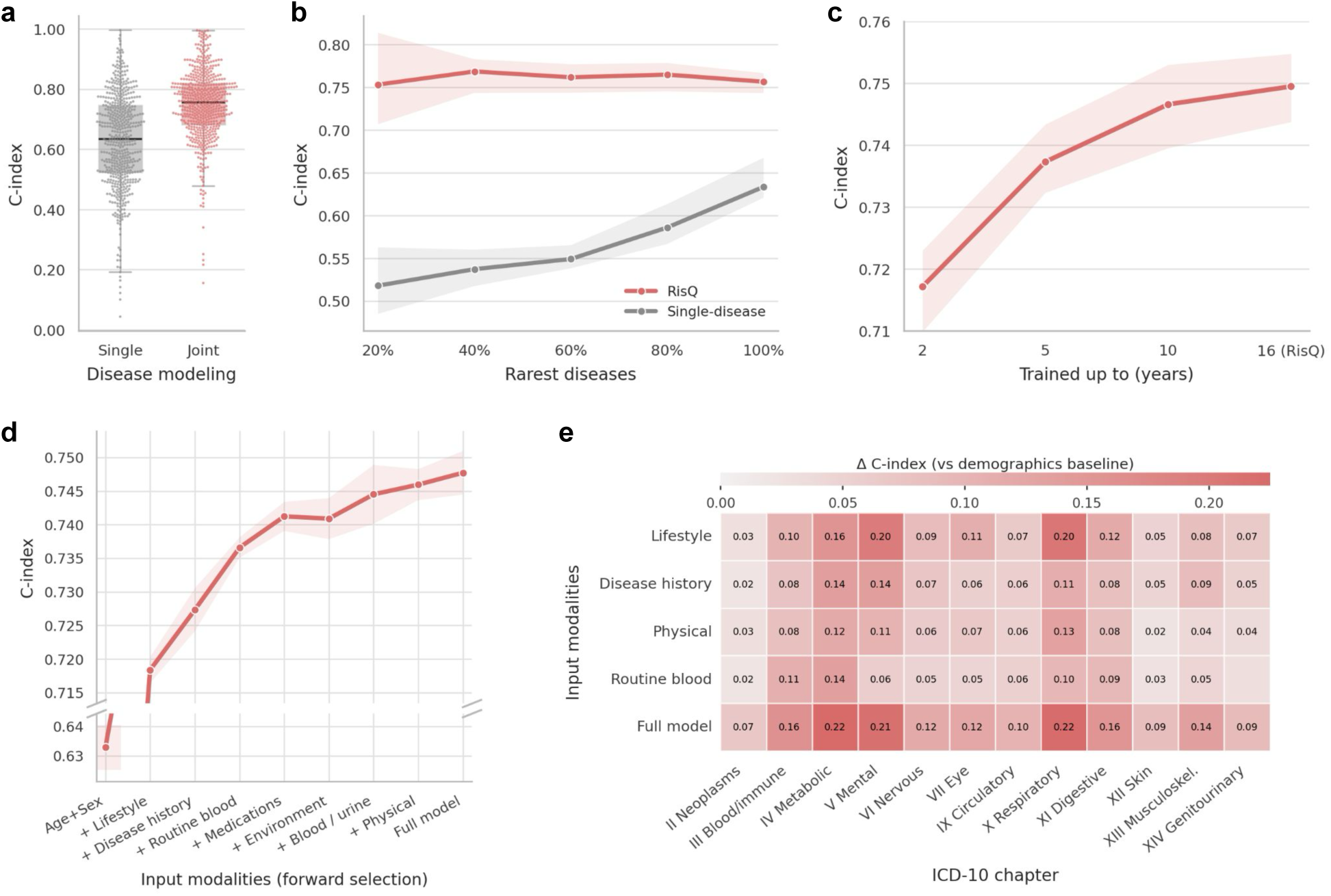
| Joint scaling across disease targets, temporal supervision, and input modalities improves risk prediction. (**a**) Multi-disease versus single-disease training using the same architecture shows best performance when diseases are modeled jointly. (**b**) Multi-disease versus single-disease training across cumulative disease sets ranked by prevalence (from the lowest 20% to all diseases) shows the largest performance gains among the rarest outcomes. (**c**) Performance increases as the amount of years included during training increases. (**d**) C-index increases as input modality groups are added using forward selection, starting from an age-and-sex baseline. (**e**) Chapter-level C-index gains relative to an age-and-sex baseline across modality configurations are plotted. The fully integrated model achieves the highest performance across ICD-10 chapters. Only significant deltas are shown. Shaded bands in (**b,c**) represent 95% bootstrap confidence intervals (2.5th–97.5th percentiles; *n* = 100 resamples). Shaded bands in (**d**) represent standard deviation across five seeds.

Finally, we assessed whether integrating diverse input modalities yields cumulative improvements in risk prediction. Variables were grouped into nine modality domains based on the UKB hierarchy and added sequentially using forward selection, beginning with an age-and-sex baseline (**Methods**). Performance improved as modalities were incorporated (Fig. 3d), and the fully integrated model achieved the highest C-index (0.748 *±* 0.003 SD). Although individual modalities contributed more strongly to specific ICD chapters, gains over the demographics baseline were observed broadly across all modality domains and no single modality matched the integrated model in any chapter (93/96 and 96/96 modality–chapter combinations significant respectively; BH-FDR *<* 0.05 each; selected deltas in Fig. 3e; full results in **Supplementary Fig. S8**).

These results show that predictive gains accumulate as we increase target diseases, time horizons, and input modalities. Together, this indicates that the learned risk structure improves with increasing information transfer when scaling all three axes.

### 2.4 Multi-scale structure of human disease risk

We next asked how the shared structure of disease risk manifests across individuals in the population. To address this question, we performed unsupervised analyses on a held-out cohort not used during model training, with consistent results across held-out folds (**Supplementary Fig. S9**, **Methods**).

At the population level, the representations were strongly organized along major demographic and physiological axes (Fig. 4a). Principal component analysis of the disease-risk scores revealed that sex was the dominant source of variation (Pearson’s *r* = 0.50 with the leading principal component), while age and body mass index (BMI) contributed additional but smaller axes of variation (maximum Pearson’s *r* = 0.14 and *r* = 0.13, respectively; **Supplementary Fig. S10**). Assigning each individual to their highest-risk disease chapter, we found that neighboring individuals in the embedding space were 3.70-fold more likely to share the same assignment than expected under random label permutations (*k* = 20, empirical *P* = 0.001, Fig. 4b), indicating that predicted disease-risk profiles exhibit local organization by disease category.

**Figure 4.**
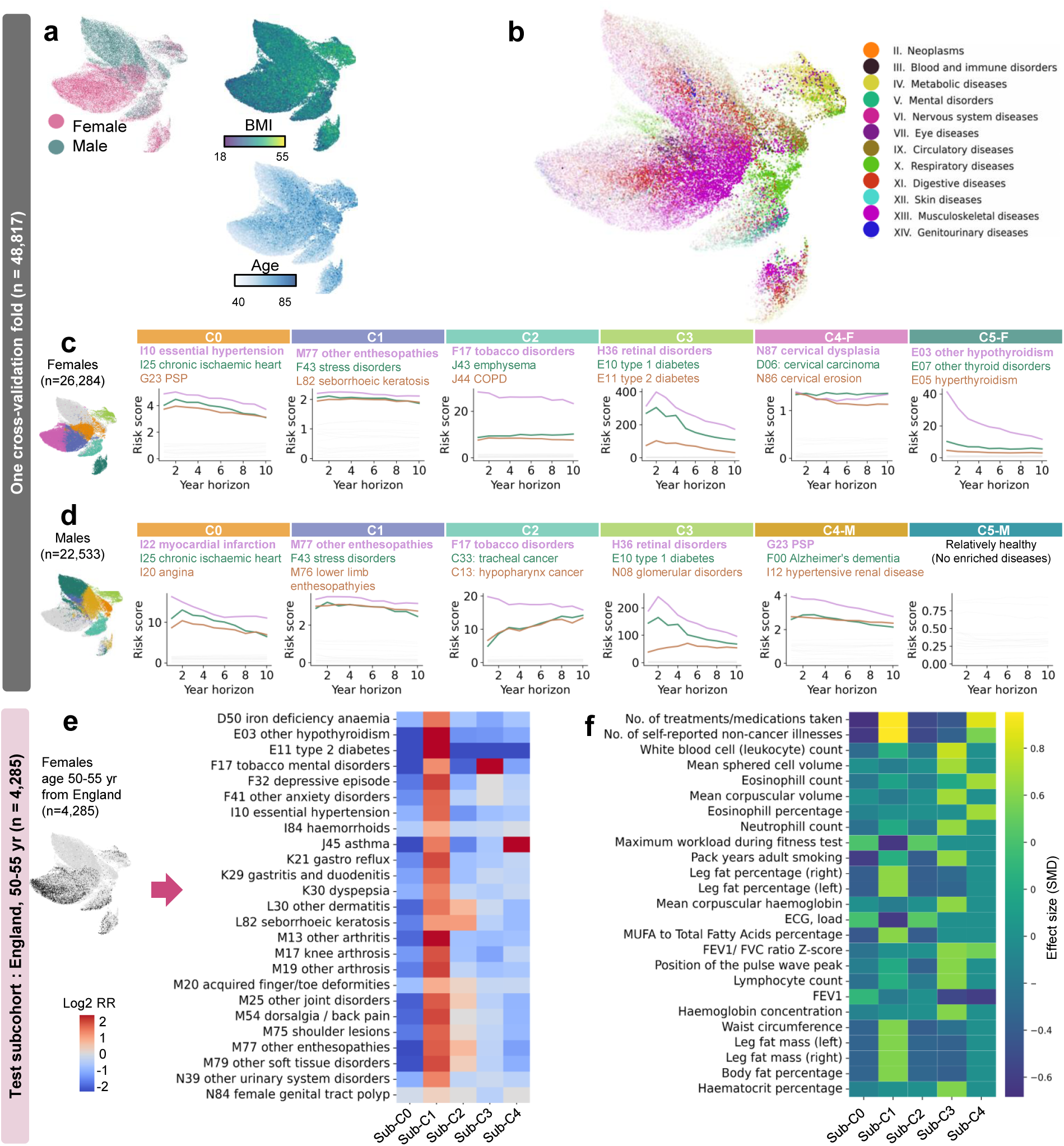
| RisQ reveals a multi-scale organization of human disease risk. (**a**) UMAP visualization of RisQ embeddings in a out-of-fold cross-validation test cohort (n=48,817), annotated by sex, body mass index (BMI), and age. (**b**) Coloring individuals by their dominant disease chapter revealed preferential localization of broad disease categories within the representation space. Each point represents one individual. Color indicates the ICD chapter of the disease with the highest mean predicted risk across prediction time points, while transparency is proportional to that disease’s mean predicted risk. (**c**) Female participants (n=26,284) were stratified into six clusters with Leiden clustering. For each cluster, risk scores are shown across prediction horizons from 1 to 10 years. Line plots show risk-ratio trajectories for the top three significantly enriched diseases in each cluster (BH-FDR*<* 0.05). (**d**) Male participants (n=22,533) were similarly stratified into six clusters, including one relatively healthy cluster (C5-M) with no enriched diseases. (**e**) Cross-disease risk profiles identified in a demographically restricted subgroup of women aged 50–55 years residing in England (n=4,285 individuals). Heatmap shows log_2_ risk ratios comparing mean 5-year predicted risks in cluster members versus non-members. Shown are diseases significantly enriched in at least one cluster (BH-FDR*<* 0.05). (**f**) Feature enrichment profiles associated with cross-disease risk clusters. Colors denote effect size (standardized mean difference, SMD). PSP: Progressive Supranuclear Palsy, COPD: Chronic Obstructive Pulmonary Disease. FEV: Forced Expiratory Volume, FVC: Forced Vital Capacity. MUFA: monounsaturated fatty acid.

We next examined how individuals partition within the risk space. Because sex was a dominant source of variation, males and females were analyzed separately to reveal finer-grained risk profiles. Within each sex, individuals partitioned into groups characterized by distinct cross-disease risk trajectories across prediction horizons (Fig. 4c**,d**; **Methods**). Several disease programs were shared across sexes, including cardiometabolic–ischaemic (C0), stress–musculoskeletal (C1), tobacco–airway (C2), and metabolic–microvascular (C3) profiles, although their relative prominence differed (Fig. 4c**,d**). Representative enrichments included chronic ischaemic heart disease (log_2_ relative risk (RR) = 1.8 and 3.1 in females and males, respectively), stress and adjustment disorders (0.99 and 1.5), tobacco-related mental disorders (4.65 and 4.09), and retinal disease (7.72 and 6.88). Additional risk profiles showed sex-differential prominence, including stronger neurodegenerative enrichment in males (C4-M) and stronger thyroid-related disease enrichment in females (C5-F), alongside a female-specific cervical neoplasms profile (C4-F; **Supplementary Dataset 2, 3**).

We next asked whether the observed risk structure captures cross-disease organization beyond broad demographic axes. To address this, we analyzed a demographically restricted subgroup defined by age, sex, and geography, focusing on women aged 50–55 years residing in England (*n* = 4,285 in this test cohort; Fig. 4e; **Methods**). Clustering identified cross-disease risk profiles broadly concordant with those observed in the population-level analysis, including relatively healthy (Sub-C0), cardiometabolic (Sub-C1), skin-musculoskeletal (Sub-C2), tobacco-associated (Sub-C3), and respiratory-enriched (Sub-C4) profiles (**Supplementary Dataset 4, 5**; **Methods**).

We then characterized the physiological, behavioral, and lifestyle signatures associated with these cross-disease risk profiles (**Methods**). We find that each risk group exhibits a distinct risk factor signature (Fig. 4f). The cardiometabolic risk profile (Sub-C1) was associated with increased BMI, elevated inflammatory markers, adverse lipid profiles, and greater treatment burden ^33^. The Sub-C2 was characterized by higher achieved workload during the UK Biobank cycle-ergometer fitness test, including higher maximum workload and higher workload recorded during ECG monitoring. The tobacco-associated risk profile (Sub-C3) showed greater smoking exposure and enrichment for leukocyte and neutrophil counts ^34,35^ (SMD = 0.67), consistent with chronic systemic inflammation linked to tobacco exposure. The respiratory-enriched risk profile (Sub-C4) exhibited elevated eosinophil counts (SMD = 0.69) and increased medication use, consistent with type 2 inflammatory airway phenotypes ^36^. Similar feature patterns were observed independently in males (**Supplementary Fig. S11**, **Supplementary Dataset 6, 7**), indicating that these cross-disease risk profiles correspond to coherent and reproducible physiological states.

Together, these analyses indicate that human disease risk exhibits multi-scale structure, spanning broad demographic axes, reproducible cross-disease risk profiles, and coherent physiological states that persist for individuals within demographically homogeneous subgroups.

### 2.5 Rare genetic variation associates with disease-risk structure

If the disease-risk structure identified by RisQ reflects biologically meaningful patterns of disease susceptibility, then naturally occurring rare genetic variation should associate with this structure. We therefore performed exome-wide gene-level loss-of-function (LoF) burden analyses across 17,984 genes using out-of-fold disease-risk embeddings generated through 10-fold cross-validation for 343,477 unrelated European UKB participants as a joint multivariate phenotype (**Methods**).

Association analysis identified 32 significant genes (Bonferroni-adjusted *P <* 0.05; Fig. 5a), which we refer to as risk-structure genes (RSGs), with calibrated permutation-based null statistics. The RSG set includes genes associated with major physiological domains, including cardiometabolic, renal, endocrine, hematologic, pigmentation, and neurologic systems (**Supplementary Dataset 8**). Over 85% had prior evidence linking them to monogenic disease, hereditary disease predisposition, protective loss-of-function effects, or somatic disease-associated variation, including well-established disease genes such as *HBB*, *GCK*, *LDLR*, and *CASR* (**Supplementary Table 1**).

**Figure 5.**
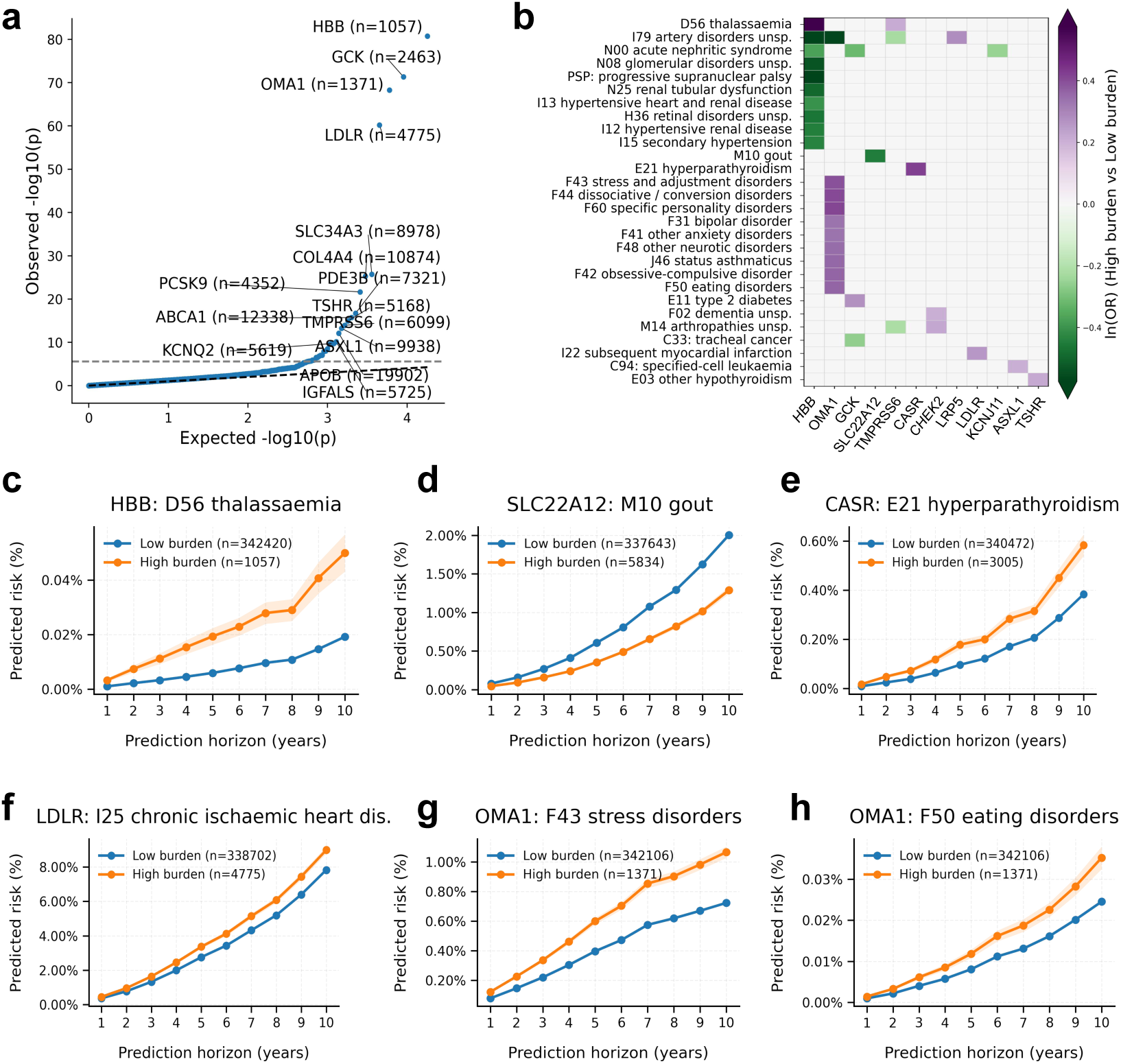
| Gene-level rare-variant burden is associated with disease-risk structure defined by RisQ. (**a**) QQ plot of gene-level LoF burden association statistics using the disease-risk embedding as a joint multivariate phenotype. Thirty-two risk-structure genes surpass Bonferroni correction (horizontal dashed line). Selected genes are labeled with high-burden counts. (**b**) Heatmap of log odds ratios (ln(OR); high versus low LoF burden) linking risk-structure genes (columns) to 10-year predicted risk across representative ICD-10 endpoints (rows). Colors denote direction and magnitude of association. (**c–f**) Predicted cumulative risk trajectories across 10-year horizons for representative canonical associations: *HBB*–thalassemia (**c**), *SLC22A12*–gout (**d**), *CASR*–hyperparathyroidism (**e**), and *LDLR*–chronic ischaemic heart disease (**f**). (**g,h**) Examples of a multi-disease association linking mitochondrial stress regulator *OMA1* to a stress-related psychiatric risk module, including stress and adjustment disorders (**g**) and eating disorders (**h**).

To link RSGs to specific disease modules, we related RSG LoF burden to predicted 10-year risk across diseases, stratifying individuals into high- and low-burden groups. This analysis revealed distinct patterns of association between genes and disease-risk profiles, with individual genes associated with both increased and decreased predicted risk across multiple conditions (Fig. 5b). Among the strongest signals, LoF burden in *HBB* was associated with increased thalassemia risk ^37^ (OR = 2.61; *P* = 4.42 *×* 10*^−^*^25^; Fig. 5c), with reduced predicted risks of several renal and hypertensive conditions (OR range 0.56–0.69), which is consistent with population-level evidence linking higher hemoglobin and hematocrit levels to elevated blood pressure and hypertension risk ^38,39^. Specific established associations included *SLC22A12*–reduced gout risk ^40^ (OR = 0.64; *P* = 3.40 *×* 10*^−^*^48^; Fig. 5d), *CASR*-hyperparathyroidism through regulation of calcium homeostasis ^41^ (OR = 1.52; *P* = 9.83 *×* 10*^−^*^42^; Fig. 5e), *LDLR*–chronic ischaemic heart disease ^42^ (OR = 1.16; *P* = 3.74 *×* 10*^−^*^23^; Fig. 5f), *TSHR*–other hypothyroidism ^43^ (OR = 1.24; *P* = 2.05 *×* 10*^−^*^24^), and *LRP5*–osteoporosis without pathological fracture ^44^ (OR = 1.06; *P* = 2.46 *×* 10*^−^*^4^; **Supplementary Dataset 9**).

The disease-risk structure also highlighted less established gene–disease links, such as the association between *OMA1* and stress-related psychiatric risk outcomes. High LoF burden was associated with elevated predicted risk for stress and adjustment disorders (OR = 1.48; *P* = 5.68 *×* 10*^−^*^18^), other anxiety disorders (OR = 1.40; *P* = 8.68 *×* 10*^−^*^15^), eating disorders (OR = 1.44; *P* = 3.32 *×* 10*^−^*^4^), obsessive-compulsive disorder (OR = 1.44; *P* = 9.72 *×* 10*^−^*^4^), bipolar disorder (OR = 1.40; *P* = 3.96 *×* 10*^−^*^3^), and dissociative/conversion disorders (OR = 1.50; *P* = 1.59 *×* 10*^−^*^2^) (Fig. 5g**,h**; **Supplementary Fig. S12**). Given that *OMA1* encodes a mitochondrial inner-membrane stress-responsive metalloprotease that regulates *OPA1* processing and mitochondrial dynamics ^45,46^, these findings suggest a candidate link between mitochondrial stress signaling and neuropsychiatric disease-risk profiles, consistent with emerging evidence implicating mitochondrial dysfunction in mood and stress-related disorders ^47^.

Together, these findings indicate that rare genetic variation associates with cross-disease risk structure, recovering established disease biology while revealing candidate gene–disease relationships for further study.

## 3 Discussion

This study provides evidence that human disease risk exhibits a shared structure spanning diseases, modalities, and time, and that leveraging this structure improves risk prediction. We demonstrated this through RisQ, a multimodal framework that jointly models diseases across time to learn a unified representation of human health. Through its query mechanism, RisQ is able to predict unseen diseases and time horizons, supporting a learnable and shared structure of disease risk. Across 588 clinician-curated diseases and up to 16 years of follow-up in the UKB, RisQ improved risk prediction relative to disease-specific gradient-boosted models and recent multi-disease foundation models by leveraging this shared structure. These findings generalized under zero-shot transfer to the independent All of Us cohort despite substantial population shift and restricted inputs.

We found that integrating increasing information across diseases, time horizons, and input modalities consistently improved risk prediction, with the largest gains observed for rare diseases and short prediction horizons. Together, these findings indicate that scaling these axes enables better information transfer through improved structure. More broadly, this supports a shift away from viewing diseases as independent endpoints toward understanding disease susceptibility as an interconnected system spanning conditions, risk factors, and time, consistent with frameworks from multimorbidity and network medicine that emphasize shared biological and phenotypic organization across diseases ^48–50^. In this view, cross-disease and shared temporal patterns reflect a common structure that can be leveraged for both prediction and analysis.

We further characterized this shared structure at multiple scales. At the population level, disease-risk profiles were shaped by major demographic variables including sex, age, and body mass index. Within demographically homogeneous subgroups, however, the same structure stratified individuals into reproducible cross-disease risk profiles associated with distinct physiological, behavioral, and lifestyle characteristics. This supports a shift towards individual-centric analysis, aligning with emerging perspectives in phenomics and multimorbidity research that emphasize studying health as a longitudinal, person-level trajectory rather than as a collection of isolated disease outcomes ^51–53^. Common structure thus organizes individuals into clusters that improve our understanding of cross-disease risk factors, which cannot be captured by studying diseases independently.

Rare genetic variation provides evidence for the biological relevance of the learned disease-risk structure. Gene-level loss-of-function burden analyses identified 32 associated genes spanning multiple physiological systems and recovered numerous established disease relationships, including *HBB* with thalassemia ^37^ and *LDLR* with ischaemic heart disease ^42^. At the same time, the analyses highlighted less characterized associations such as *OMA1* and a stress-related psychiatric risk profile. This is consistent with extensive evidence for pleiotropy and shared genetic effects across complex traits, and supports the interpretation of disease-risk structure as a population-scale representation linking molecular variation to patterns of multimorbidity ^54,55^. In this framework, shared genetic influences may contribute to coordinated patterns of disease susceptibility across conditions which can be recovered by analyzing the shared latent structure.

These findings should be interpreted in light of several limitations. The UKB is a healthy-volunteer cohort enriched for European ancestry and middle-to-older age at enrollment, potentially limiting generalizability. Recorded diagnoses reflect healthcare access and coding practices; accordingly, the model predicts recorded disease rather than biological disease onset ^56^. Although the external All of Us evaluation helps mitigate these concerns, our analyses emphasize discrimination rather than clinical utility, and translation would require additional evaluation using decision-focused metrics such as net benefit ^57,58^. Finally, associations between rare genetic variation and the disease-risk structure do not establish causal mechanisms and may be influenced by confounding and collider biases inherent to observational healthcare data ^59,60^.

Taken together, our findings support studying disease susceptibility as an interconnected system rather than as a collection of independent disease outcomes. More broadly, they suggest the need for future health systems that are increasingly organized around continuously updated representations of individuals, integrating information across ever expanding measurements of lifestyle, biology, and environment. Looking forward, such dynamic representations of health could provide a common framework for anticipating disease, monitoring health trajectories, and uncovering biological mechanisms across the human lifespan.

## 4 Methods

### 4.1 Data

#### 4.1.1 UK Biobank

##### Cohort and study design

The UK Biobank ^19^ served as the primary training and in-distribution evaluation cohort. Baseline (instance 0) assessment data and linked longitudinal health-record follow-up were analyzed in 488,170 participants with baseline variables and longitudinal outcome fields. Participants were recruited across the UK between 2006 and 2010 and were aged 37–73 years at baseline. The index date was defined as the date of attendance at the baseline assessment centre. Structured variables were derived from baseline assessment measurements and questionnaires. Longitudinal diagnoses were obtained from linked primary care (GP), hospital inpatient admissions, cancer registry, death registry, and UK Biobank-curated first-occurrence records. 46.03% of participants possessed linked GP records.

##### Input variables and disease outcomes

Model inputs comprised 1,370 structured variables derived from 1,153 UK Biobank (UKB) assessment fields, together with diagnosis history and medications (**Supplementary Dataset 1**, **10** & **11**). Structured variables comprised demographics (age and sex; fields 31 and 21003); questionnaire and cognitive-function items; physical measures; blood and urine assays; local environment variables; polygenic risk scores and related genetic summaries; and self-reported medical history and current medications (field 20003). Continuous and integer fields were Gaussian-quantile normalized using parameters estimated from the training set. Categorical fields were integer-coded with missing values retained. Structured variables, diagnosis-history tokens, and medication-history tokens yielded 4,463 input features. Disease outcomes comprised first-occurrence diagnoses (category 1712), neoplastic diseases derived from linked inpatient and cancer-registry records, and algorithmically defined outcomes (**Supplementary Table 2**). Outcomes were defined by the first recorded post-baseline occurrence date, and participants with prevalent disease were excluded for that target. The full disease panel comprised 1,246 prognosis targets. Targets were collapsed to three-character ICD-10 prefixes and full disease name as defined in the UKB cohort and encoded using BioLORD (**Supplementary Fig. S2**). Primary analyses were restricted to 588 clinician-curated targets, excluding congenital, pregnancy-related, perinatal, injury and external-cause codes. Results for all 1,246 targets are reported in **Supplementary Fig. S2**. A list of all diseases can be found in **Supplementary Dataset 1**.

##### Outcome definition, censoring, and cohort splits

Follow-up was right-censored at the earliest of death (field 40000), nondiabetic hypoglycaemic coma (field 130716), or administrative censoring on May 31, 2022. Placeholder event dates flagged by UK Biobank were treated as missing. Participants with GP records were partitioned into training, validation, and test sets using a 70:15:15 split and a fixed random seed. Validation and test sets were restricted to participants with linked GP records to minimize false negatives, whereas the training set additionally included participants without GP linkage. This yielded 420,760 participants in the training set and 33,705 participants each in the validation and test sets. All UKB benchmark models were trained and evaluated on this common split. Details about cohort characteristics can be found in **Supplementary Table 3**.

#### 4.1.2 All of Us

##### Cohort and diagnosis harmonization

External validation used the All of Us Research Program Registered Tier Curated Data Repository (version 8, released February 2025), analyzed within the All of Us Researcher Workbench ^61^. Model inputs comprised age, sex at birth, body-mass index (BMI), drinking behavior, smoking behavior, and diagnosis history. Age was computed as (index date *−* birth datetime)*/*365.25 years. Sex at birth was obtained from the participant record. BMI was derived from LOINC 39156-5 using the measurement closest to and preceding the index date within a one-year window. Drinking and smoking behaviors were derived from All of Us survey responses and mapped to the corresponding UKB variables (Drinking and smoking mappings can be found in **Supplementary Table 4** and **Supplementary Table 5**, respectively). Disease outcomes were harmonized between All of Us and UKB by mapping OMOP condition concepts to the corresponding UKB ICD-10 targets using OMOP standard concepts. The study cohort comprised 257,538 participants with at least one diagnosis mapping to the UKB target set. Details about cohort characteristics can be found in **Supplementary Table 6**.

##### Evaluation

All models were evaluated using records available up to the date of survey completion, which served as the index date. For each participant and target prefix, diagnosis history was defined by the earliest condition-occurrence date for that prefix. Targets without a faithful All of Us equivalent were excluded from the external validation (354 targets retained). For each target, outcomes were defined by the first diagnosis occurring after the index date; participants with the target diagnosed on or before the index date were excluded as prevalent. Follow-up was right-censored at the earliest of death or the administrative cut-off of October 1, 2023. Participants whose index date occurred after the administrative cut-off were excluded. All models were evaluated without retraining, fine-tuning, or recalibration. RisQ, Delphi-2M and XGBoost comparators were evaluated at annual prediction horizons from 1 to 5 years using the same evaluation framework as in UKB. Prediction was limited to 5 years as this was the maximum time range supported by the data while maintaining case counts.

### 4.2 RisQ architecture and training

#### RisQ architecture

RisQ is a transformer-based model for disease-risk prediction from multimodal data. Structured covariates, diagnosis histories, and medications were represented as modality-specific input tokens and encoded into a latent health representation using a transformer encoder. Disease-risk predictions were generated by conditioning this representation on a query specifying a target disease and prediction horizon, enabling risk estimation across multiple diseases and time horizons. The architecture comprises multimodal input representations, a transformer encoder that produces a latent representation, and a query-based disease risk decoder that generates disease- and horizon-specific predictions.

#### Input tokenization

Structured covariates were embedded using feature-specific tokenizers in the style of the FT-Transformer ^20^, with missing values omitted. Diagnosis history and medications were represented as variable-length token sequences, derived by projecting pretrained BioLORD ^21^ embeddings of the corresponding clinical concepts into the RisQ model dimension. A UMAP of diagnosis embeddings can be found in **Supplementary Fig. S2**. These diagnosis history tokens were additionally augmented with sinusoidal encodings of time since diagnosis.

#### Health representation

The transformer encoder generated latent representations of individuals from the input token sequence. A fixed number of learnable health tokens were appended to the input sequence prior to encoding. Information from modality-specific input tokens was aggregated into the latent tokens through self-attention, yielding a compact representation used for downstream risk prediction. The encoder used a stack of transformer layers following the FT-Transformer ^20^ architecture, details can be found in the code repository.

#### Query-based prognosis decoder

Disease-risk predictions were generated from an individual’s health representation using a cross-attention decoder. For each target disease, a disease-specific query was constructed from the same pretrained ICD embedding used for diagnosis-history tokenization and augmented with a sinusoidal encoding of the queried prediction horizon. Disease–horizon queries attended to the latent health representation to generate one prediction logit per target and horizon. No self-attention was applied among prognosis queries, and targets were decoded independently conditional on the same health representation.

#### Horizon-conditional training objective

Training labels were defined with respect to sampled prediction horizons. For positive participant–target pairs, horizons were sampled from a Gaussian distribution centered on the observed event time with a standard deviation of 12 months. For non-event pairs, horizons were sampled uniformly over the available observation time. Given a sampled horizon, labels were defined by whether the event occurred before that horizon. RisQ therefore learned horizon-conditional event probabilities rather than directly regressing time to event.

#### Model training and inference

RisQ was trained using binary cross-entropy over all valid participant–target pairs per step in a batch of 128 subjects using gradient accumulation over 4 steps, resulting in a batch size of 512. Optimization used AdamW with learning rate 3 *×* 10*^−^*^3^, weight decay 1 *×* 10*^−^*^5^, and linear learning-rate warm-up over 10 epochs. Modality dropout was applied with probability 0.6 by randomly masking predefined modality groups, adapted from the UKB data category structure with selected variables reassigned to semantically aligned groups (e.g. parental disease history moved to the genetics group; full group definitions in **Supplementary Dataset 10**), while preserving a protected age-and-sex core feature set. RisQ shown in Fig. 2 is an ensemble of 15 independently seeded models. During inference, the model was queried at fixed 1-year prediction horizons to generate disease-risk predictions. Predicted disease-risk was constrained to be non-decreasing.

### 4.3 Baseline models

We compared RisQ to gradient-boosted trees^23^ (in both classification and survival formulations), the TabPFN ^22^ tabular foundation model, the Delphi-2M ^18^ generative disease-history transformer, and a set of established clinical risk scores. Experimental setup was identical for all models.

#### XGBoost

We implemented classification and survival gradient-boosted tree baselines using the XGBoost library. Diagnosis history was encoded as a multi-hot ICD vector with temporal values, storing the number of months between baseline and the most recent pre-baseline occurrence of each ICD code (0 indicating no prior occurrence). Medications were encoded as a boolean multi-hot vector. Missing values were not imputed. Binary classifiers were trained on 1-, 2-, 5-, and 10-year horizons. Cox survival models were trained over all non-censored data points. Both a default and optimized variant of each model was evaluated on each task. Optimized models were selected from Bayesian hyperparameter searches of 20 trials per (disease, horizon)-pair for classification models and 20 trials per disease for survival models. Hyperparameter sweep configurations and default values are reported in **Supplementary Table 7**. This resulted in 3,550 disease–horizon classifiers and 1,176 disease-specific survival models, and over 47,260 hyperparameter configurations tested.

#### TabPFN

TabPFNv3^22^ is a tabular foundation model that performs Bayesian-inspired in-context learning over structured data, enabling prediction without dataset-specific training. We evaluated TabPFNv3 on the 1-, 2-, 5-, and 10-year horizons using a train set of 100,000 target-stratified individuals drawn from the training split for each endpoint. The reported performance is obtained from the identical held-out test set. Even with subsampling, average runtime per disease was about an hour resulting in a total evaluation time of approximately 1,764 GPU hours.

#### Delphi-2M

Delphi-2M ^18^ is a generative transformer trained on tokenized longitudinal disease-history sequences with minimal structured data. To ensure comparability with our experimental setting, we retrained Delphi-2M on the same UKB split common to all other models, following the original Delphi specification. As a sanity check, we first reproduced the originally reported Delphi-2M performance before adapting the model to our fixed-horizon evaluation setting.

Because Delphi-2M has been designed as a next-event predictor rather than a risk estimator, we adapt it to risk estimation using the following evaluation protocol. For each individual, we define an anchor as their assessment center visit and truncate its input sequence to history observed up to that anchor. Delphi-2M can then produce disease-specific scores from this pre-anchor history, and performance is evaluated prospectively for incident disease within fixed horizons starting from the anchor. This anchoring approach produced the strongest and most stable results among all adaptation strategies we tested (**Supplementary Fig. S13**).

#### Clinical risk scores

Clinical risk scores are expert-derived predictive models that estimate an individual’s disease risk using a small set of predefined input variables and fixed weighting schemes. Clinical score implementations were adapted to the available UKB baseline fields; participants missing any required covariate for a given score were excluded from that score-specific analysis. Each endpoint was defined as the earliest incident post-baseline across the respective ICD codes targeted by the score. Score specific subject eligibility restrictions such as age ranges or variable-conditioned subsets were respected. Granular adaptation and mapping steps can be found in our code repository.

For cardiovascular disease, we evaluated three risk scores: the Framingham 2008 general cardiovascular disease score ^25^, the 2013 ASCVD pooled cohort equations ^26^ and SCORE2^27^. For type 2 diabetes, we evaluated FINDRISC ^28^ and an ADA Diabetes Risk Test-style score ^29^. For lung cancer, we computed the PLCOm2012 risk score ^30^. Kidney failure was evaluated using the Kidney Failure Risk Equation ^31^.

### 4.4 Prediction evaluation

#### Evaluation metrics

The primary metric for classification was the area under the receiver-operating-characteristic curve (AUC), computed for each disease and horizon pair from the model’s risk scores and the binary incident-case labels. For survival analysis we calculated the concordance index (C-index). We report Antolini’s time-dependent C-index, computed from the predicted survival curves. Unless noted otherwise, all reported metrics are the macro or mean over all diseases. Of the 588 clinician-curated disease targets, 545 had sufficient case counts across splits (*≥* 10 positive cases in training, *≥* 1 in each of validation and test) and were retained for evaluation; the remaining 43 were excluded. For AUC panels, confidence intervals were obtained by non-parametric subject-level bootstrap resampling with 100 resamples, discarding any bootstrap sample containing only a single outcome class. For C-index confidence intervals, two schemes were used: disease-level resampling of the macro-median for the prevalence-stratified analysis (Fig. 3b), and subject-level resampling of the macro C-index for the zero-shot chapter generalization and temporal training-cap analyses (Fig. 1d; Fig. 3c), each with 100 resamples.

#### Zero-shot chapter generalization

To evaluate generalization to unseen disease chapters, we adopted a leave-one-chapter-out (LOCO) protocol. For each of the 12 ICD-10 chapters represented in the 588-target panel, a separate RisQ model was trained with all targets from that chapter withheld from supervision. At test time, held-out targets were predicted by querying the corresponding chapter-withheld model. Predictions across the 12 models were stitched to cover all 569 held-out ICD targets (excluding 19 algorithmically defined outcomes without direct ICD-10 chapter assignment; **Supplementary Table 2**). Macro and per-chapter C-indices were computed on these stitched predictions using subject-level bootstrap resampling (100 resamples; 2.5th–97.5th percentiles). A fully supervised RisQ model and an age-and-sex-only RisQ model were evaluated on the same 569 targets as references.

#### Natural-language querying

To evaluate generalization of the query mechanism to unseen query formats, a single fully supervised RisQ model (trained on all 588 targets with standard BioLORD ICD embeddings) was queried at test time using free-text natural-language disease descriptions instead (**Supplementary Fig. S2c**). No chapter withholding or stitching was performed. Performance was reported on the same 569 ICD-chapter-assignable targets for direct comparability with the chapter generalization results.

#### Temporal extrapolation

To evaluate generalization to unseen time horizons, independent RisQ models were trained with observation caps of 2, 5, and 10 years, restricting sampled training horizons to within each cap. All models were evaluated at annual prediction horizons from one to ten years. For caps below ten years, this includes horizons beyond the training cap. AUC at each horizon was computed using subject-level bootstrap resampling (100 resamples; 2.5th–97.5th percentiles).

#### Statistical comparison of models

Unless stated otherwise, model comparisons used matched per-disease performance differences as the statistical unit. The precise number of matched/available diseases is provided in **Supplementary Table 8**. For each disease, the median performance metric across bootstrap replicates ^62^ was computed for each model, and two-sided Wilcoxon signed-rank tests ^63^ were applied to the paired per-disease differences.

This framework was used for zero-shot chapter generalization (Fig. 1d), discrimination comparisons (Fig. 2a**,b**), low-prevalence subgroup analyses (Fig. 2d**,e**), external validation AUC comparisons (Fig. 2j), multi- versus single-disease ablations (Fig. 3a**,b**), temporal training-cap comparisons (Fig. 3c), and multimodal integration analyses (Fig. 3e). For multimodal integration, tests were performed within ICD-10 chapters using one-sample Wilcoxon tests on per-disease C-index differences.

Multiple-testing correction was applied as appropriate: Bonferroni correction ^64^ across all matched diseases in all 24 comparisons across 4 horizons (1, 2, 5, 10) *×* 6 baselines (Fig. 2a); Bonferroni correction across 6 comparisons across 3 horizons (1, 2, 5) *×* 2 baselines (Fig. 2j); no correction for the single low-prevalence comparison per horizon; Benjamini–Hochberg false discovery rate (FDR) correction across the five cumulative prevalence buckets for Fig. 3b, across the three cap comparisons for Fig. 3c, and across modality–chapter combinations for Fig. 3e.

### 4.5 Disease-risk structure analysis

#### Disease-risk representations

Disease-risk representations were derived from model-predicted disease-risk trajectories in the UKB out-of-fold test set (*n* = 48,817). Clustering analyses were restricted to a single cross-validation fold to ensure that all representations were embedded in a common prediction space. For each individual, predicted probabilities across 588 diseases and 1–10 year horizons were aggregated into a participant-by-(disease *×* horizon) matrix, followed by principal component analysis (PCA) retaining the top 8 components (84.2% variance explained). Associations between embedding dimensions and demographic variables (sex, age, BMI) were quantified using Pearson correlation (**Supplementary Fig. S10**). For visualization, embeddings were projected into two dimensions using UMAP (Fig. 4a**,b**). To assess local organization, each individual was assigned a dominant ICD-10 chapter based on the chapter-wise highest mean predicted risk. Neighbourhood enrichment was quantified as the fraction of *k* = 20 nearest neighbours sharing the same label, compared to random permutations to assess significance.

#### Sex-specific cluster analysis

Disease-risk groups were identified independently in males and females by applying the Leiden community-detection algorithm ^65^ to the 8-dimensional disease-risk representations (n_neighbors=40, resolution=0.2). The resolution parameter was selected based on stability analysis, balancing cluster reproducibility and interpretability (**Supplementary Fig. S9**). To characterize disease-risk patterns, predicted risks were compared between individuals in each cluster and all remaining individuals of the same sex using risk ratios (RR), with standard errors estimated via the delta method. Statistical significance was assessed using *χ*^2^ tests with Benjamini–Hochberg correction (BH-FDR*<* 0.05). For visualization, the top three significantly enriched diseases per cluster were reported (Fig. 4c). Correspondence between male and female disease-risk groups was assessed using the Hungarian algorithm to maximize profile similarity, with concordance quantified by Jaccard overlap of the top-*k* (*k* = 10) enriched diseases.

#### Disease-risk structure in a demographically restricted population

Disease-risk structure was additionally analyzed in a restricted subgroup of women aged 50–55 years in England (*n*=4,285), focusing on a 5-year prediction horizon for 25 diseases with mean predicted risk *≥* 1% (**Supplementary Dataset 4**). Principal component analysis of the participant-by-disease risk matrix retained the top 10 components (88% variance explained), followed by Leiden clustering using the same parameters as in the sex-specific analysis. Disease-risk patterns were characterized using the same enrichment framework, retaining diseases significant at BH-FDR*<* 0.05. A cluster-by-disease enrichment matrix was constructed from all significant associations (Fig. 4e), and clusters were assigned descriptive labels based on their enrichment profiles.

#### Risk factor profiles

To identify features associated with disease-risk groups in the demographically restricted population analysis, we compared feature distributions between in-cluster and out-of-cluster individuals using a one-versus-rest design within the analysis subset. For continuous features, enrichment was quantified using the standardized mean difference (SMD), defined as the difference in mean feature value between groups divided by the pooled standard deviation. For categorical features, global association was assessed using Fisher’s exact test (binary) or Pearson’s *χ*^2^ test (multi-level), and effect sizes were summarized using log_2_ relative risk (RR), with Haldane–Anscombe correction applied where necessary; for multi-level features, the level with the largest absolute log_2_ RR was retained as the representative effect. Multiple testing across all features was controlled using the Benjamini–Hochberg procedure (BH-FDR *<* 0.05), and significant features were ranked by absolute effect size (SMD for continuous features; log_2_ RR for categorical features). For visualization, we report the union of the top 25 significantly enriched or depleted features per cluster (Fig. 4f). Equivalent analyses were performed independently in males and females to assess reproducibility of cluster-associated feature profiles (**Supplementary Fig. S11**).

#### Cluster stability analyses

To assess the stability of participant assignments, Leiden clustering was repeated across different resolution parameters ranging from 0.15 to 1.0. Resulting cluster assignments were compared with the reference clustering (resolution 0.2) using the adjusted Rand index (ARI) and normalized mutual information (NMI) on the same participants. Resolution 0.2 is chosen to balance biological interpretability with stability, without the over-fragmentation seen at high resolutions. To evaluate the reproducibility of disease-risk groups across independent participant subsets, clustering was performed separately on the disjoint test sets from each outer cross-validation fold (disjoint test sets per fold). Clusters were matched between runs by risk-profile similarity using the Hungarian algorithm, with correspondence scored by Spearman correlation of cluster-wise log_2_ risk-ratio profiles and Jaccard overlap of top-ranked enriched diseases (topk = 10). The same profile-based matching was used to compare male and female disease-risk groups and to compare population-level (sex-stratified) clustering with the restricted England, women aged 50–55 years subgroup, because these analyses involve non-overlapping participants and sample-level ARI/NMI are not applicable. For fold comparisons, one fold was designated as the reference and matched clusters were summarized by median profile similarity and disease recurrence in the union top-10 enrichment set. Cluster stability results are reported in **Supplementary Fig. S9**.

### 4.6 Genetic analyses

#### Gene-level burden association analysis

We tested for associations between gene-level loss-of-function (LoF) burden and RisQ disease-risk embeddings to identify genetic factors associated with shared disease-risk structure. Gene-level LoF burden scores were computed from whole-exome sequencing data using the pretrained DeepRVAT model ^66^. Disease-risk embeddings were derived by applying principal component analysis (PCA) to out-of-fold prediction logits across 588 diseases and 1–10 year prediction horizons, retaining the top 40 components (>90% variance explained). To avoid information leakage, logits were generated using 10-fold cross-validation such that each individual was scored only by models not trained on that individual. Prior to PCA, logits were residualized for age, sex, genetic ancestry principal components, and fold membership, and resulting components were transformed using a rank-based inverse normal transformation ^67,68^. Associations between gene-level burden and the 40-dimensional embedding were tested using a multivariate GWAS framework (MT-GWAS ^69^), treating the embedding as a joint phenotype. Analyses were performed in 343,477 unrelated European UKB participants across 17,984 genes with at least one rare variant (MAF *<* 0.1%) observed in at least 10 individuals. Covariates included age, sex, age^2^, sex-by-age interactions, genotyping array, 40 genetic principal components, and 36 common-variant polygenic risk scores **Supplementary Table 9**.

#### Disease-level characterization of significant genes

Genes reaching Bonferroni significance in the gene burden association analysis were further characterized through disease-specific risk profiles and longitudinal trajectories. For each significant gene, we compared 10-year RisQ-predicted disease risks between higher-burden and lower-burden individuals, defined using a DeepRVAT score threshold of 0.75. Differences in predicted risk were quantified using odds ratios computed from mean predicted probabilities in the two groups, OR = [*p̅*_high_*/*(1 − *p̅*_high_)]*/*[*p̅*_low_*/*(1 − *p̅*_low_)]. Standard errors of ln(OR) were approximated using the delta method, and association statistics were computed as *z* = ln(OR)*/*SE(ln(OR)), with two-sided *P* values derived from *z*^2^ under a *χ*^2^ distribution. For each gene, associated diseases were identified at BH-FDR*<* 0.05 using the Benjamini–Hochberg procedure. Gene–disease association heatmaps were restricted to pairs with at least 30 observed cases and to associations satisfying BH-FDR*<* 0.05 and *|* ln(OR)*| ≥* 0.2. For visualization, the 10 diseases with the largest absolute log odds ratios per gene were retained. For selected significant gene–disease pairs, we additionally visualized predicted disease-risk trajectories for higher- and lower-burden individuals across 1–10 year prediction horizons.

### 4.7 AI usage

The authors used OpenAI ChatGPT and Codex/Code, Anthropic Claude and Claude Code, and Google Gemini to assist with software development and quality assurance, and to provide language and editorial support during manuscript preparation. All generated code and text were reviewed, tested, and edited by the authors, who are solely responsible for the final content of this manuscript.

## Data Availability

The data used in this study are available through the UK Biobank and the All of Us Research Program. Researchers may apply for access through the respective access procedures.

https://allofus.nih.gov

https://www.ukbiobank.ac.uk

## 5 Code and Data Availability

All code, including data pre-processing, model training, evaluation, and analysis code will be made available at https://github.com/RisQ-Lab/RisQ. Access to data from the UKB and All of Us cohorts can be applied for on the respective project websites.

## 6 Author Information

These authors contributed equally: Paul Hager, Benedikt Roth, Niklas Bühler, Diyuan Lu

These authors jointly supervised this work: Daniel Rueckert, Fabian Theis, Francesco Paolo Casale

## Contributions

PH, FPC, FT and DR conceptualized the study. PH, DL and FPC designed the RisQ framework. PH led implementation. BR, NB and DL performed model evaluation and downstream analyses. JHB and LS contributed to validation and data processing. RKR, ER, JP, MDF, EC, JAS and LA contributed to interpretation and provided feedback. DR, FT and FPC supervised the research. PH, BR, NB, DL and FPC wrote the manuscript. All authors contributed to revising the manuscript and approved the final version.

## 7 Acknowledgments

We thank Na Cai for feedback on the manuscript. This research has been conducted using the UK Biobank Resource under Application Number 87065. The authors would like to acknowledge the participants of the All of Us Research Program, without whom this research would not be possible, and the program’s funding partners, which include the National Institutes of Health. This study used data from the National Institutes of Health’s All of Us Research Program’s Controlled Tier Dataset version 8 (CDRv8), available to authorized users on the Researcher Workbench (Workspace Identifier: aou-rw-09d045b8). Benedikt Roth and Liubov Shilova acknowledge the support of the research school Munich School for Data Science (MUDS). Benedikt Roth, Jamison H. Burks, and Eljas Roellin received funding from the Innovative Medicines Initiative 2 Joint Undertaking (JU) under grant agreement No. 101007873 (UNITE4TB). The JU receives support from the European Union’s Horizon 2020 research and innovation programme and EFPIA, Deutsches Zentrum für Infektionsforschung e.V. (DZIF), and Ludwig-Maximilians-Universität München (LMU). EFPIA/Associated Partners contribute 50% of the funding, whereas the contribution of DZIF and LMU University Hospital Munich has been granted by the German Federal Ministry of Education and Research. Diyuan Lu was supported by the Chan Zuckerberg Initiative (CZI) through the AI Residency Program. Liubov Shilova was funded by Friedrich-Alexander-University Erlangen-Nuremberg. Maxime Di Folco has received funding from the French National Research Agency (ANR) as part of the France 2030 plan under grant agreement ANR-22-EXES-0013. Francesco Paolo Casale was funded by the Free State of Bavaria’s High-Tech Agenda through the Institute of AI for Health (AIH). This work was partially funded by ERC Grant Deep4MI (Grant No. 884622) and DFG Grant No. 566800501.

## A Supplementary Figures

**Figure S1.**
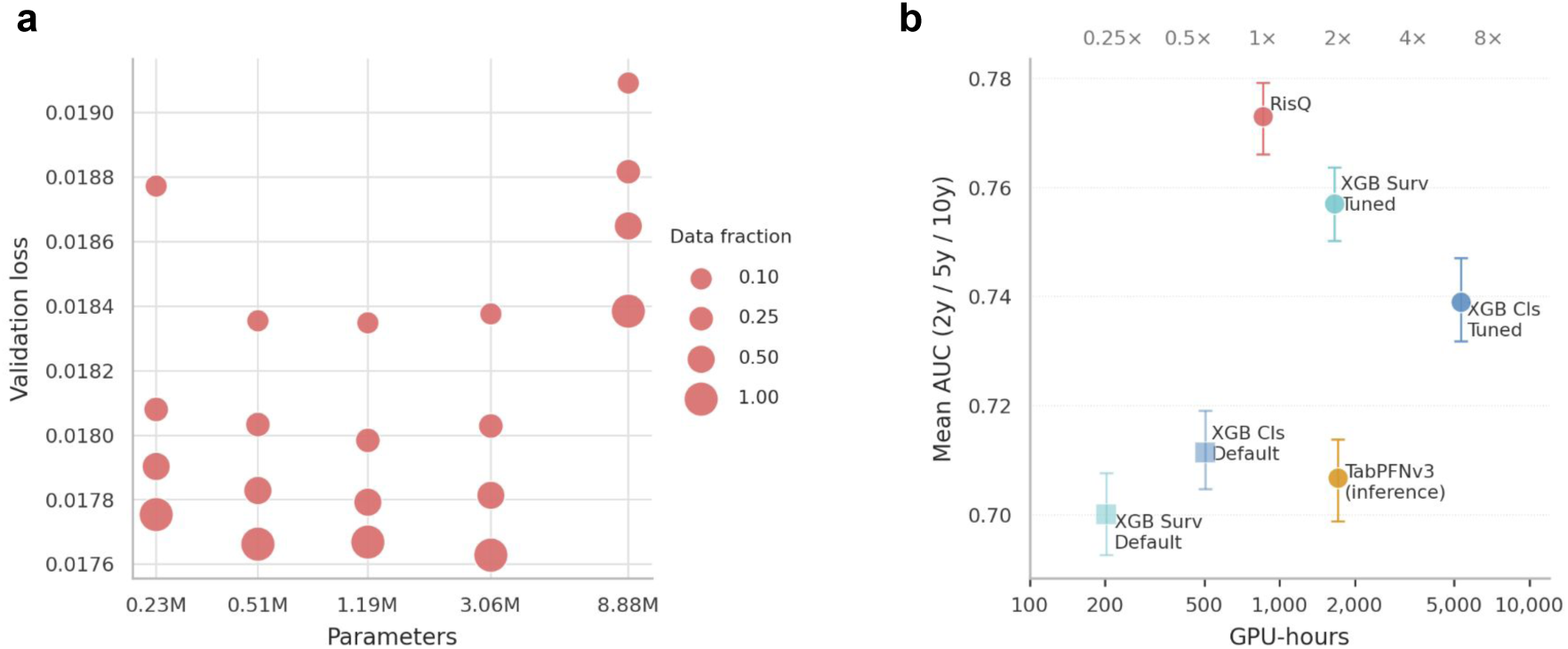
| Scaling analysis and compute efficiency. (**a**) Validation loss versus trainable parameters for RisQ model and data-fraction scaling; marker area denotes training-data fraction (10%, 25%, 50%, and 100%). (**b**) Pareto plot of mean macro AUC (averaged across 2-, 5-, and 10-year prediction horizons) versus total GPU-hours. All models were executed on identical GPU hardware to ensure directly comparable compute costs. Error bars indicate 95% bootstrap confidence intervals. The top axis shows compute relative to RisQ. Square markers denote default hyperparameters; circles denote tuned models. TabPFNv3 costs reflect inference only (no training or hyperparameter tuning). RisQ training cost covers all 588 targets and prediction horizons in a single run, whereas baseline compute scales per target and per horizon.

**Figure S2.**
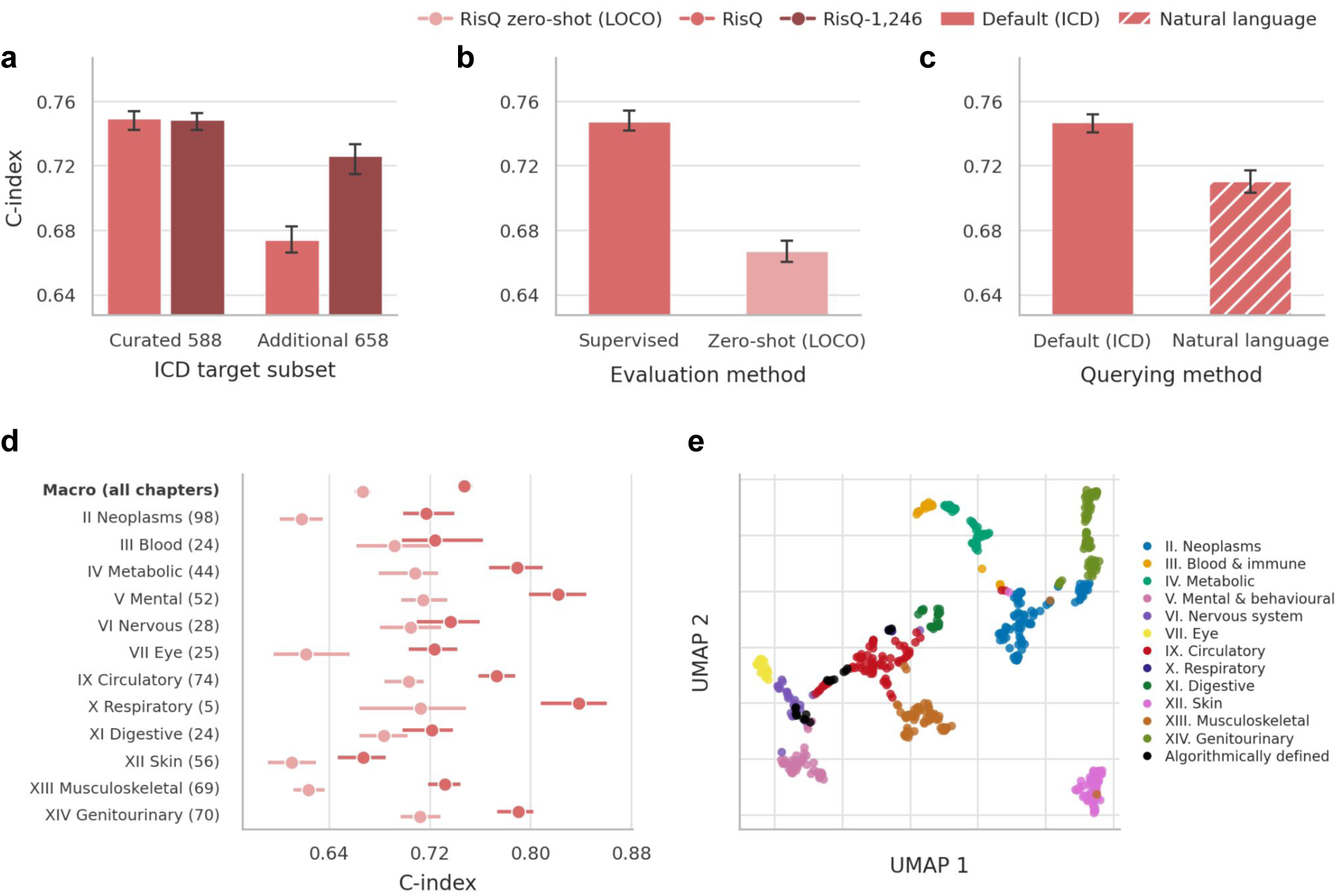
| Evaluation on all 1,246 ICD targets and zero-shot generalization. (**a**) Macro C-index on the 588 clinician-curated ICD targets used for RisQ training and the additional 658 targets not supervised in that model, comparing RisQ (trained on the curated 588) and RisQ-1,246 (trained on all 1,246 targets). RisQ matched RisQ-1,246 on curated targets but underperformed on the additional 658 zero-shot stratum; full-panel training (RisQ-1,246) substantially reduced this gap. (**b**) Macro C-index on 569 ICD chapter hold-out targets (19 algorithmically defined outcomes without direct ICD-10 chapter assignment excluded; **Supplementary Table 2**), comparing supervised RisQ with RisQ zero-shot (LOCO). (**c**) Macro C-index on the same 569 targets using default BioLORD ICD query embeddings versus free-text natural-language queries (e.g. *What is the risk of acute myocardial infarction?*). Natural-language queries yield lower macro C-index than ICD embeddings. (**d**) Per-chapter C-index on the 569 chapter hold-out targets, comparing RisQ and RisQ zero-shot (LOCO); macro row shown at top. (**e**) Two-dimensional UMAP visualization of 768-dimensional BioLORD-2023 embeddings for all ICD-coded prognosis targets. Points are colored by ICD-10 chapter; the 19 algorithmically defined outcomes are shown in black, illustrating that ICD chapter structure is preserved in the embedding space. Error bars indicate 95% confidence intervals from participant-level bootstrap resampling (2.5th–97.5th percentiles; *n* = 100 resamples) for C-index panels.

**Figure S3.**
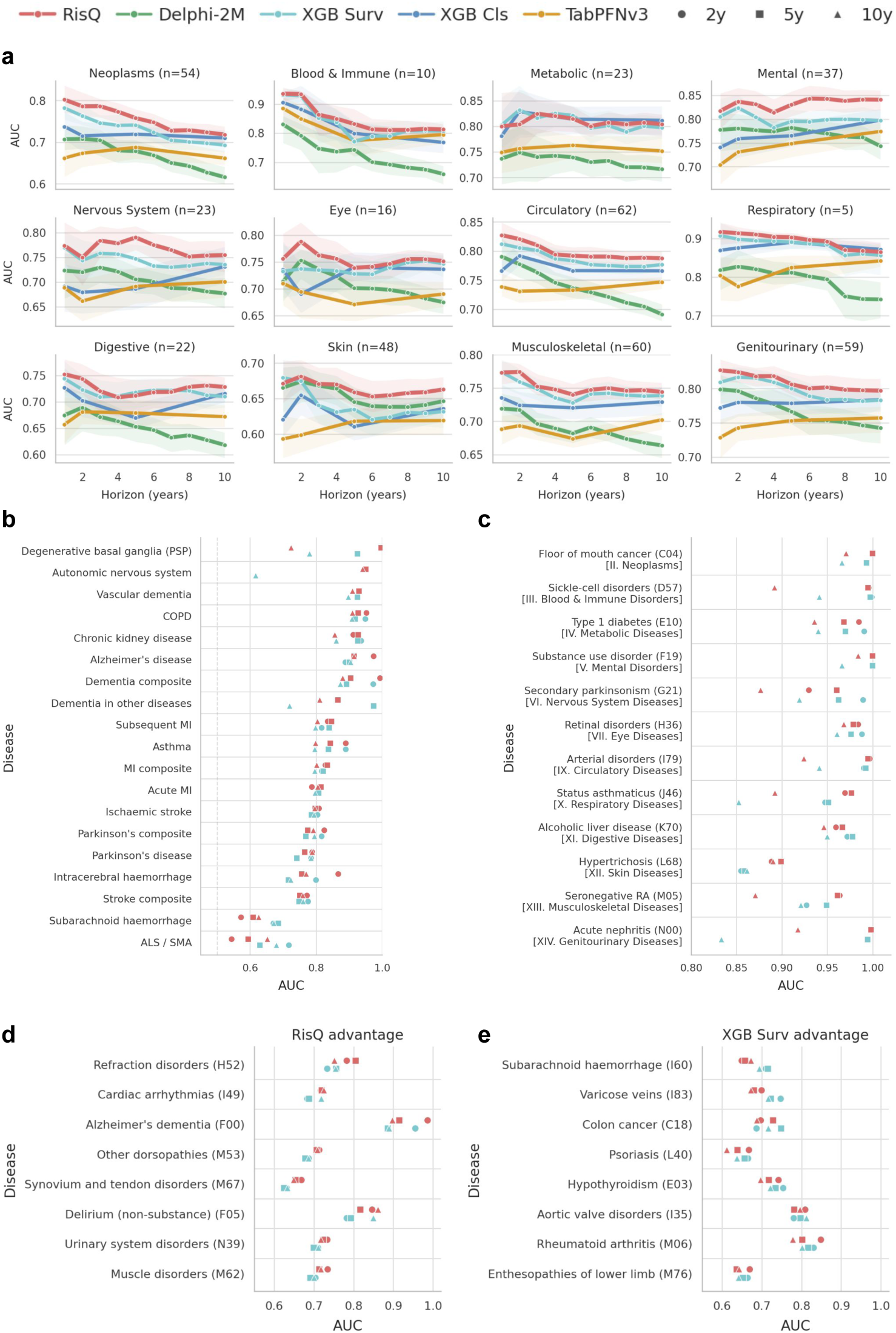
| Disease-level prediction performance across ICD-10 chapters and endpoints. (**a**) Mean AUC as a function of prediction horizon (1–10 years) for all five models, stratified by ICD-10 chapter; *n* denotes the number of matched targets at the 5-year horizon. Shaded regions indicate 95% bootstrap confidence intervals. (**b**) AUC for 19 algorithmically defined UK Biobank endpoints (RisQ and XGB Surv), sorted by RisQ 5-year AUC in descending order. (**c**) Best-performing disease per ICD-10 chapter based on RisQ 5-year AUC, comparing RisQ and XGB Surv across 2-, 5-, and 10-year horizons. (**d**) Diseases with the largest RisQ advantage over XGB Surv at 5 years (ΔAUC = RisQ *−* XGB Surv), restricted to diseases with *≥* 300 incident test-set events and ranked by ΔAUC in descending order. (**e**) As in (**d**), but for diseases where XGB Surv outperforms RisQ. All metrics are median AUC with 95% bootstrap confidence intervals (2.5th–97.5th percentiles; *n* = 100 resamples).

**Figure S4.**
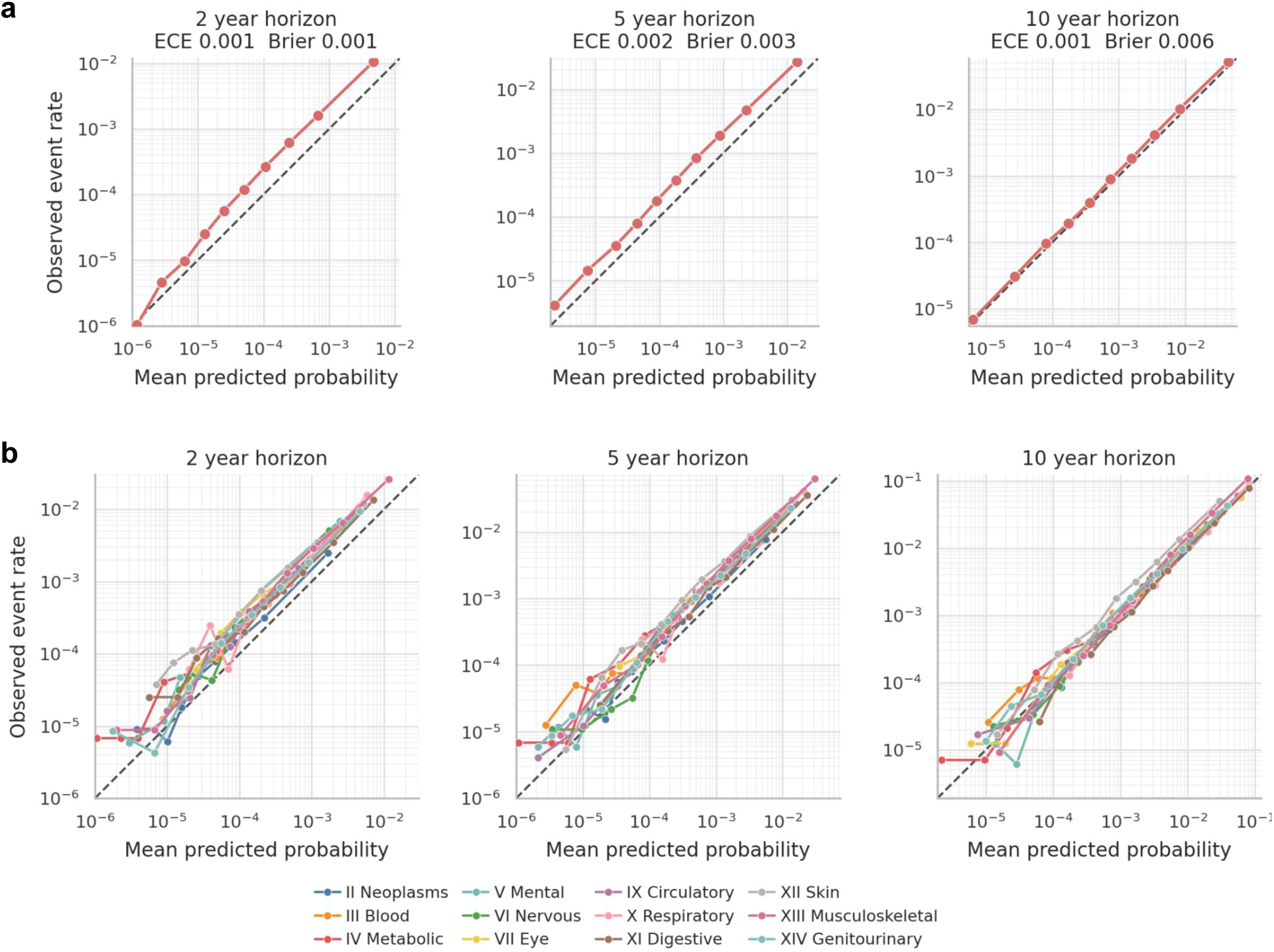
| Calibration of predicted risks at 2-, 5-, and 10-year horizons across 588 diseases. Calibration curves show observed versus mean predicted event probabilities using pooled subject–disease pairs with censoring-adjusted estimation and 10 quantile bins. The dashed diagonal indicates perfect calibration. (**a**) Overall calibration across all diseases (ECE and Brier score shown). (**b**) Calibration stratified by ICD-10 chapter.

**Figure S5.**
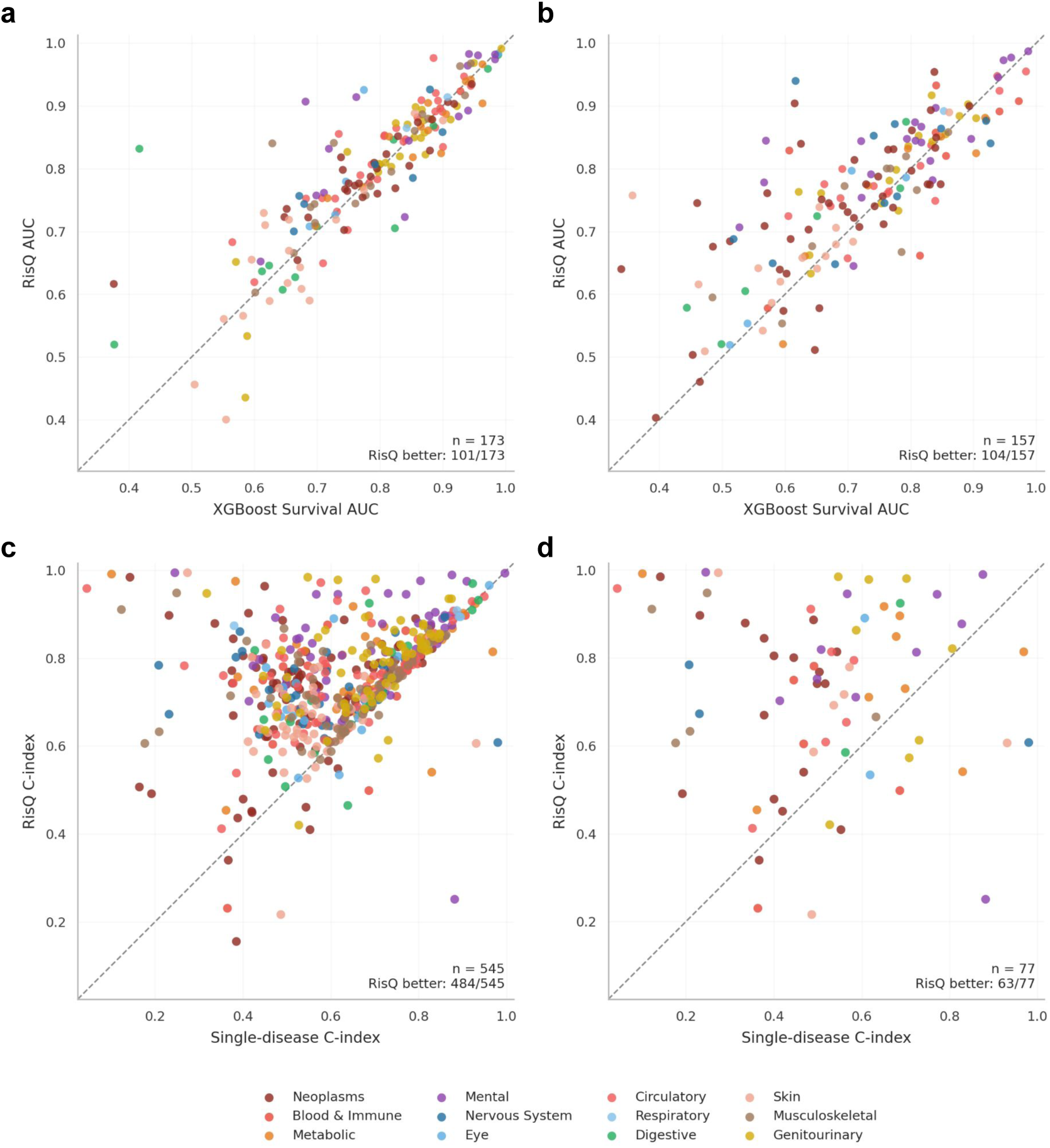
| Per-disease discrimination: low-prevalence performance and joint multi-disease training. (**a**) Per-disease AUC of RisQ versus optimized XGBoost Survival at the 2-year prediction horizon for diseases with test-set incidence 0.01–0.1%. (**b**) As in (**a**), at the 10-year prediction horizon. Points above the diagonal favor RisQ in all panels. (**c**) C-index of RisQ versus disease-specific single-disease models across all diseases, colored by ICD-10 chapter. The dashed diagonal indicates equal performance; points above the line favor joint training. RisQ outperforms the corresponding single-disease model for 484 out of 545 diseases with at least one positive test case. (**d**) As in (**c**), restricted to the 20% lowest-prevalence diseases (*n* = 77). RisQ outperforms single-disease models for 63 of 77 diseases.

**Figure S6.**
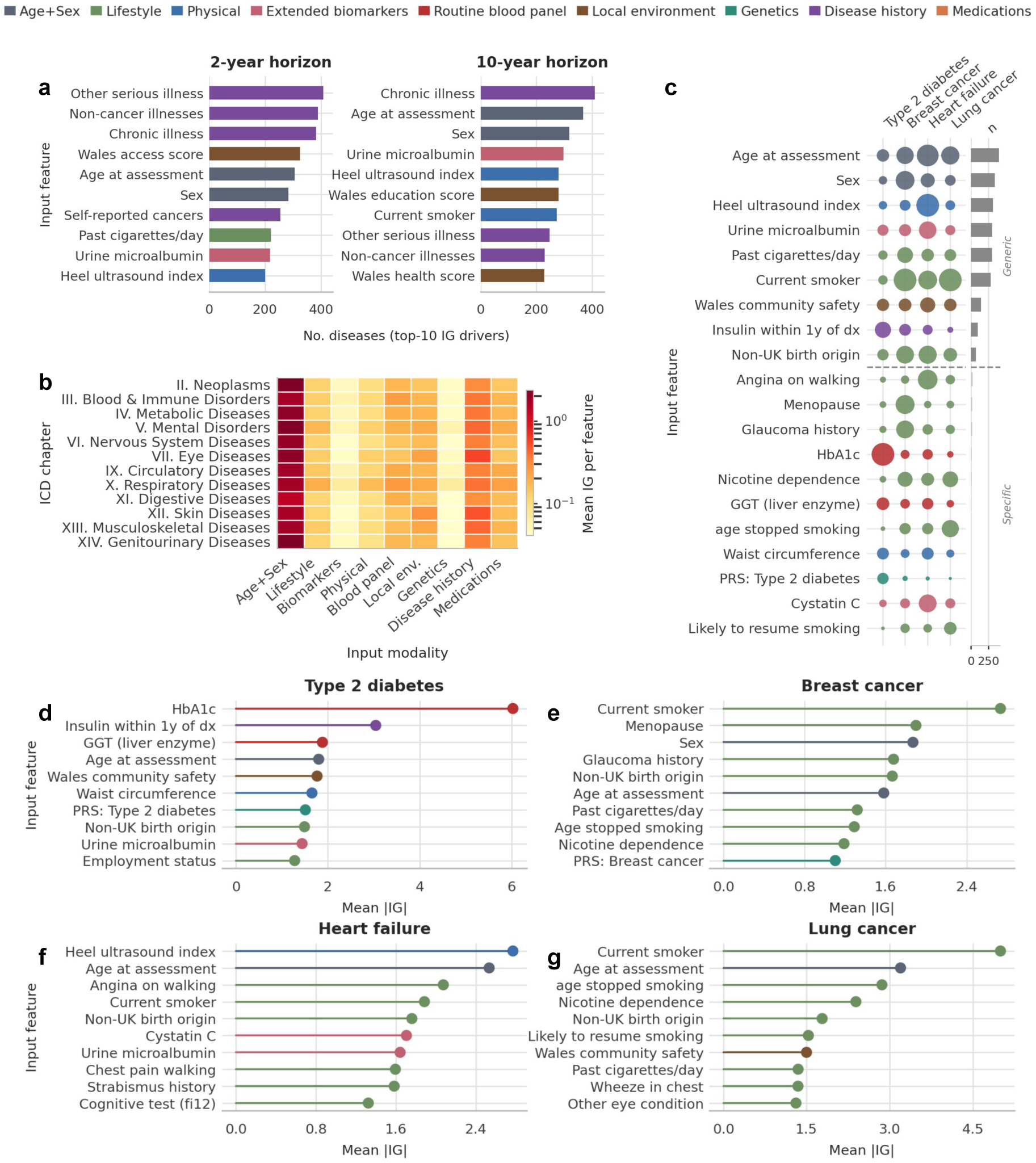
| Integrated gradient (IG) feature attribution across 588 disease outcomes. (**a**) Top recurrent IG drivers at the 2-year (left) and 10-year (right) horizons. Bar length indicates the number of diseases for which a feature ranks among the top-10 IG contributors. Bars are coloured by input modality (legend). (**b**) Mean IG aggregated by ICD-10 chapter (rows) and input modality (columns), averaged across 2- and 10-year horizons and normalised by the number of features per modality. Colour intensity reflects mean IG on a log scale. (**c**) Disease-specificity matrix for four exemplar diseases. Rows represent features from the union of per-disease top-7 IG drivers. Dot size is proportional to normalised absolute IG and colour denotes input modality. Features below the dashed line are disease-specific (ranked among the global top-10 IG drivers in *≤*10% of diseases), whereas features above are broadly recurrent. The right bar indicates the total number of diseases in which each feature appears among the global top-10 IG drivers. (**d–g**) Mean absolute IG for the top-10 features for Type 2 diabetes (**d**), Breast cancer (**e**), Heart failure (**f**), and Lung cancer (**g**). Features are ranked by absolute IG magnitude and values are averaged across the 2- and 10-year horizons.

**Figure S7.**
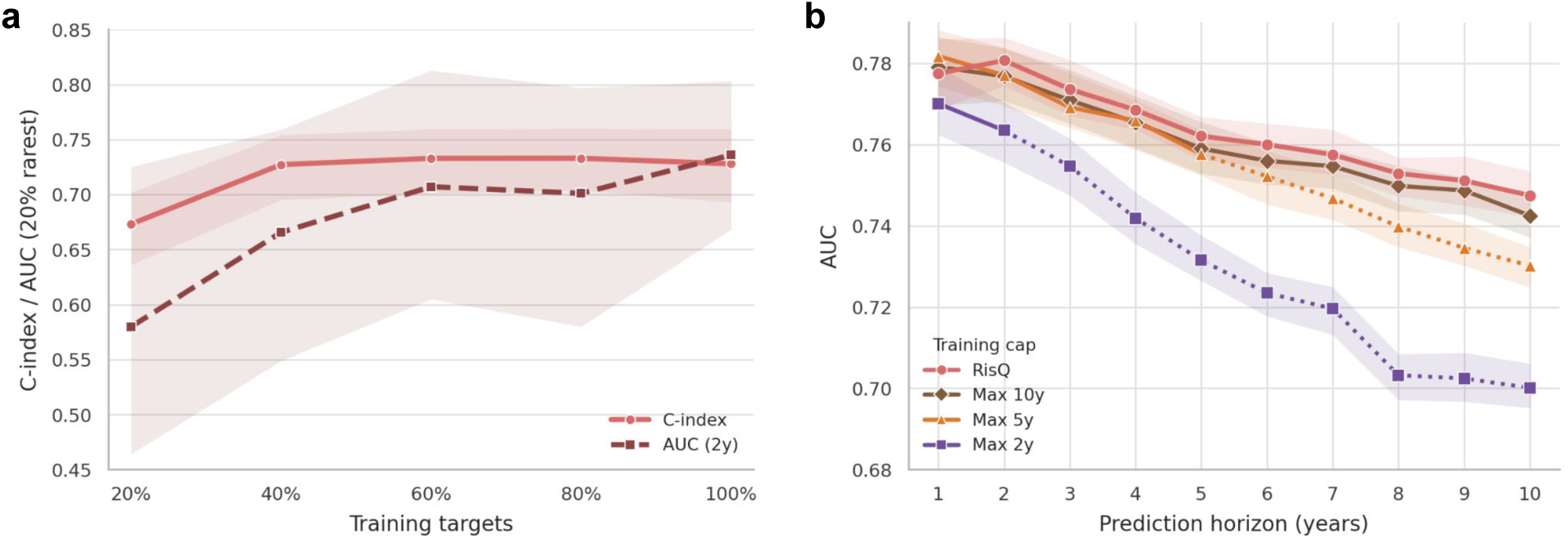
| Supplementary analysis of rare-disease performance and temporal training horizons. (**a**) Macro C-index (solid) and 2-year AUC (dashed) on the 20% rarest ICD targets (test-set prevalence), as a function of the fraction of training targets included in supervision. Models were trained on cumulative prevalence slices from the rarest targets upward (20%, 40%, 60%, 80%) and compared with fully supervised RisQ (100%). Performance on rare endpoints improved with broader supervision. (**b**) Macro AUC across all 588 clinician-curated targets versus prediction horizon (1–10 years), comparing uncapped RisQ with diagnosis-centered training restricted to maximum observation windows of 2, 5, or 10 years (*Max 2y*, *Max 5y*, *Max 10y*). All models were evaluated at every horizon; capped models are shown with solid lines up to their training cap and dotted lines beyond it. Shorter training caps reduced long-horizon discrimination, with the largest deficit for *Max 2y*. Shaded bands and error bars indicate 95% confidence intervals from individual-level bootstrap resampling (2.5th–97.5th percentiles; *n* = 100 resamples).

**Figure S8.**
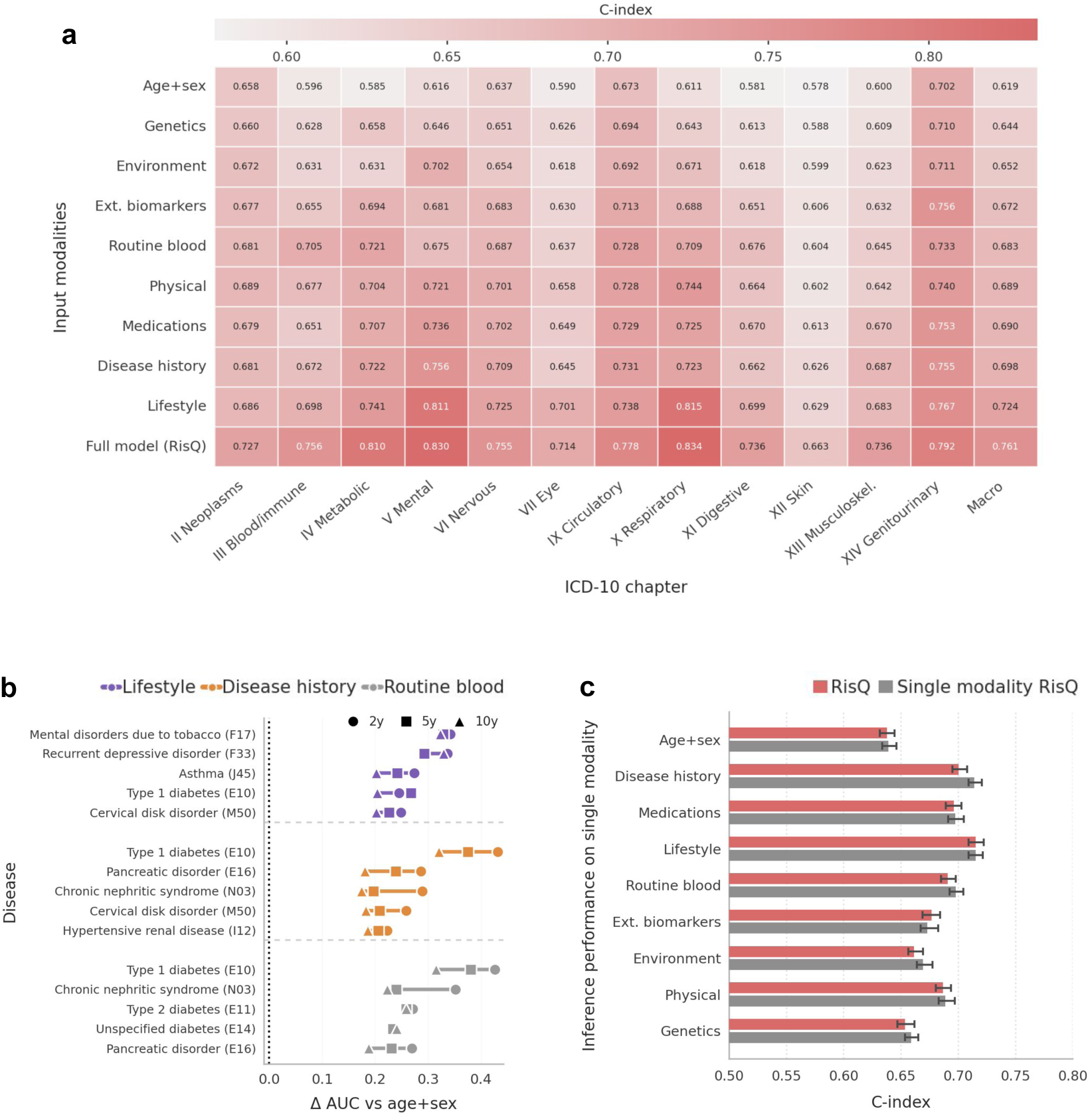
| Ablation of input modalities and robustness to restricted inference. (**a**) Test-set C-index across ICD-10 chapters for models using individual modality groups, alongside the age-and-sex baseline and the full multimodal model. Modality contributions vary by disease chapter, indicating domain-specific gains, whereas the full model achieves the highest overall performance and the best performance within each chapter. (**b**) Disease-level view of modality ablations for selected modality groups. For each modality, the five diseases with the largest improvement in discrimination relative to the age-and-sex baseline are shown as ΔAUC, restricted to outcomes with at least 20 positive test-set cases at 2-, 5-, and 10-year horizons. The largest gains occur in clinically coherent disease–modality pairs, supporting the medical specificity of the modality effects observed in (**a**). (**c**) Inference-time ablation. The full multimodal model, trained with modality dropout, was evaluated using only a single modality (plus age and sex) and compared with models trained directly on the same restricted inputs. Performance is similar across modalities, indicating that the full model remains effective under missing-modality settings without retraining dedicated modality-specific models.

**Figure S9.**
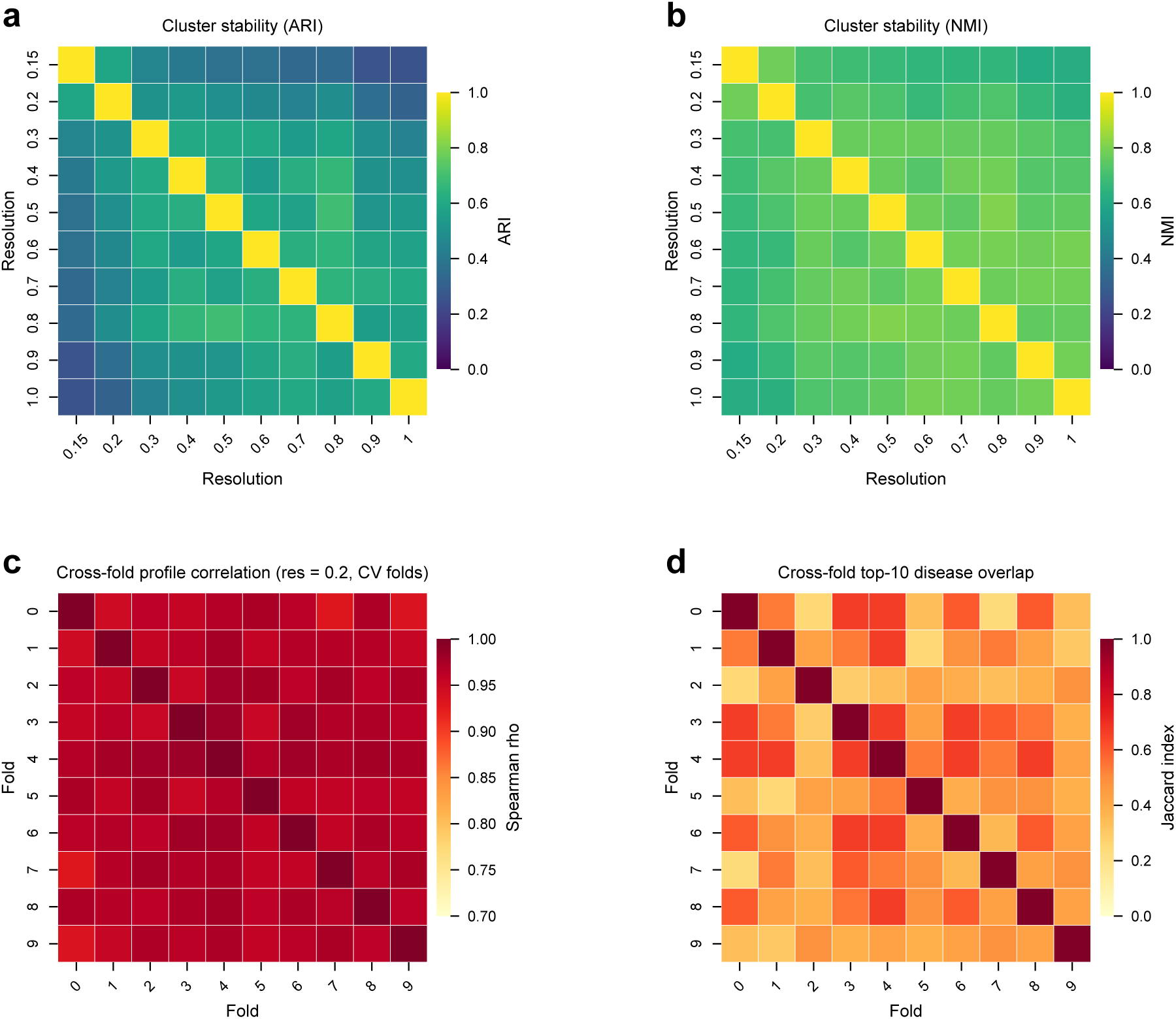
| Cluster and disease-program stability across resolution and cross-validation folds. (**a, b**), Pairwise cluster stability among female participants in outer cross-validation fold 2 (n = 26,284), assessed by comparing sample-level Leiden assignments across ten resolutions (0.15–1.0; 8 PCs). (**a**), Adjusted Rand Index (ARI). (**b**), Normalized Mutual Information (NMI). Brighter colours indicate greater agreement between partitions; both metrics show moderate-to-high stability across resolutions. We selected resolution 0.2 for downstream analysis because it yields a compact, interpretable partition while retaining high cross-resolution agreement with finer clusterings (NMI 0.67–0.72 versus 0.3–0.8; 0.78 versus 0.15), indicating that higher resolutions mainly subdivide existing programs rather than define qualitatively new ones. Resolution 0.2 therefore balances biological interpretability with stability, without the over-fragmentation seen at resolutions *≥* 0.9. (**c**), Cross-fold reproducibility of disease enrichment profiles at resolution 0.2, quantified as the median Spearman correlation (*ρ*) of log_2_ risk-ratio vectors between Hungarian-matched cluster pairs from ten outer CV folds. (**d**), Cross-fold overlap of top-10 enriched diseases for matched clusters, summarized by the median Jaccard index. High profile correlation together with moderate top-disease overlap indicates that disease programs are consistently recovered across independent test folds despite modest variation in exact disease membership.

**Figure S10.**
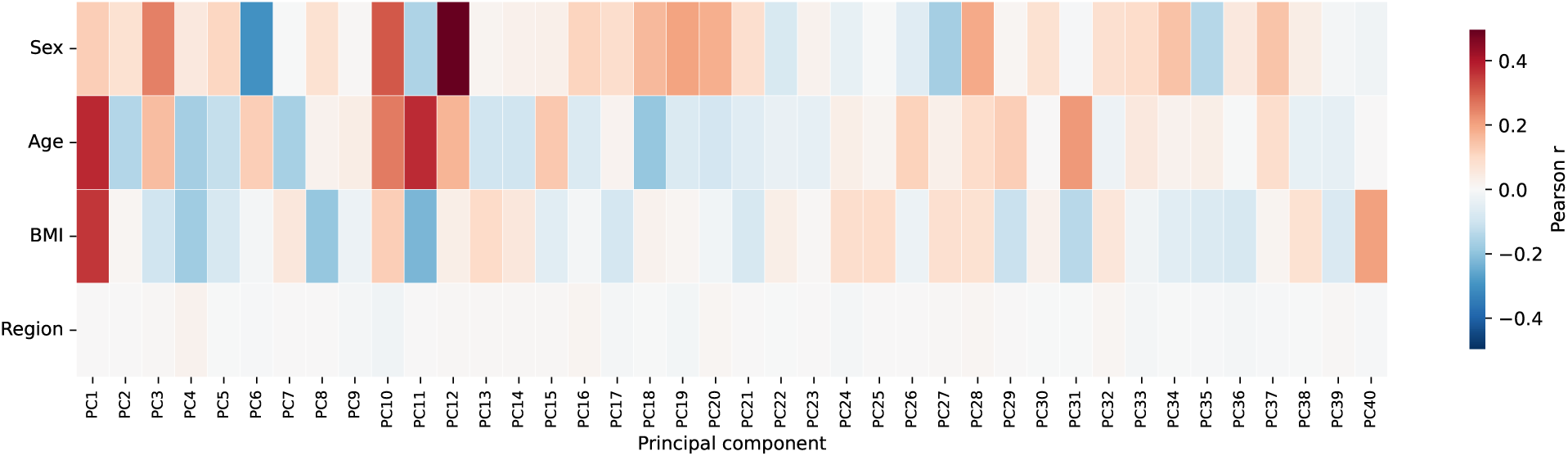
| Association of demographic covariates with disease-risk principal components. Heatmap of Pearson *r* between each retained PC (x-axis) and sex, age, BMI, and region (y-axis) in the held-out test cohort (whole fold; *n* = 48,817).

**Figure S11.**
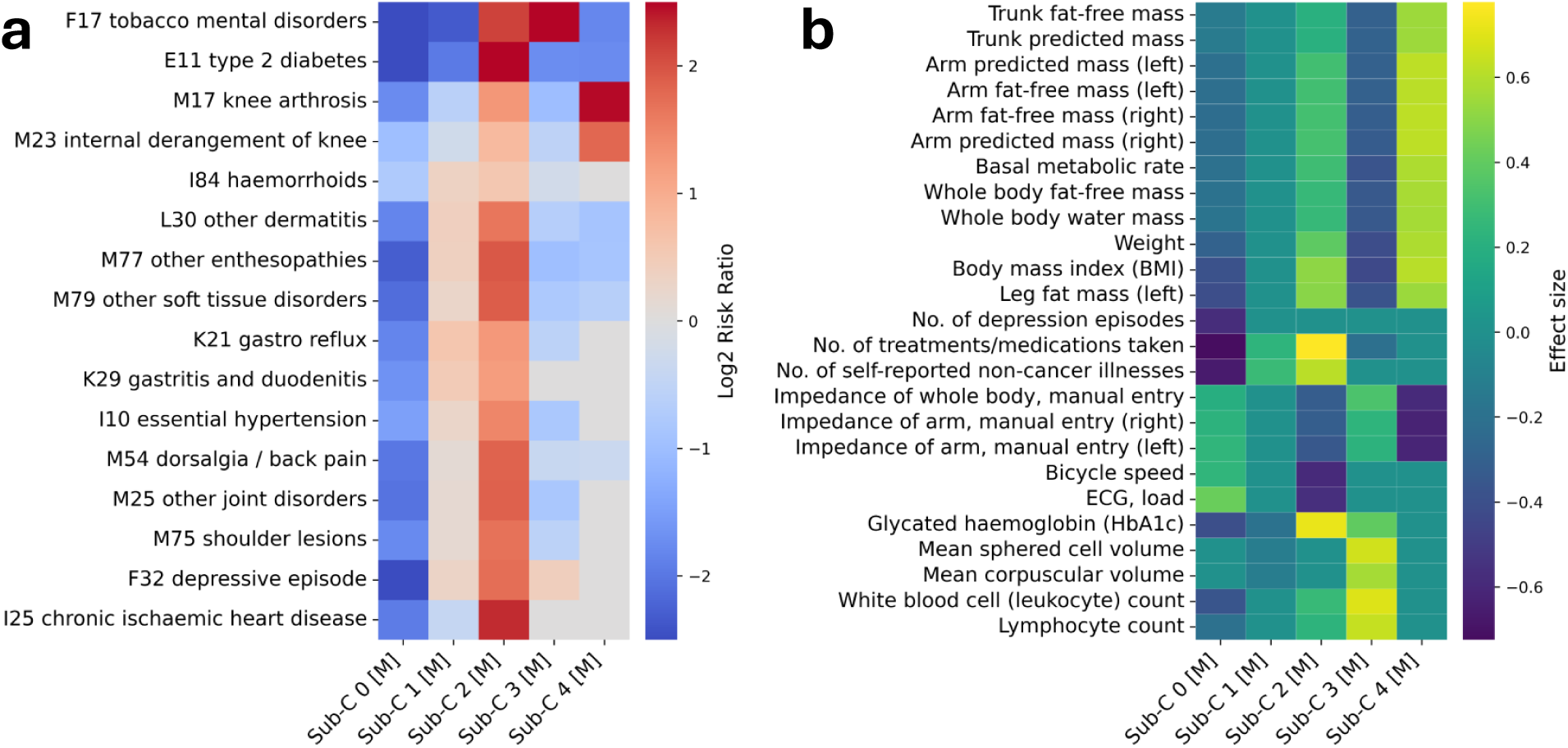
| Male clustering recapitulates cross-disease risk and feature profiles observed in restricted females cohort. (**a**) Heatmap of representative disease-risk enrichments across male-specific clusters, quantified as the (log_2_) risk ratio comparing mean predicted risk among cluster members with that among all other male participants. (**b**) Enrichment of selected continuous physiological and clinical features, quantified using standardized mean differences. The male analysis recovered broadly concordant disease programs and associated feature patterns, including tobacco-associated mental-health, cardiometabolic, respiratory, and general high-risk profiles. Notably, the general high-risk cluster (Sub-C2) showed lower LDL-related lipid measures despite broad disease-risk enrichment, together with greater use of cholesterol-lowering medication, consistent with a treatment-modified lipid profile.

**Figure S12.**
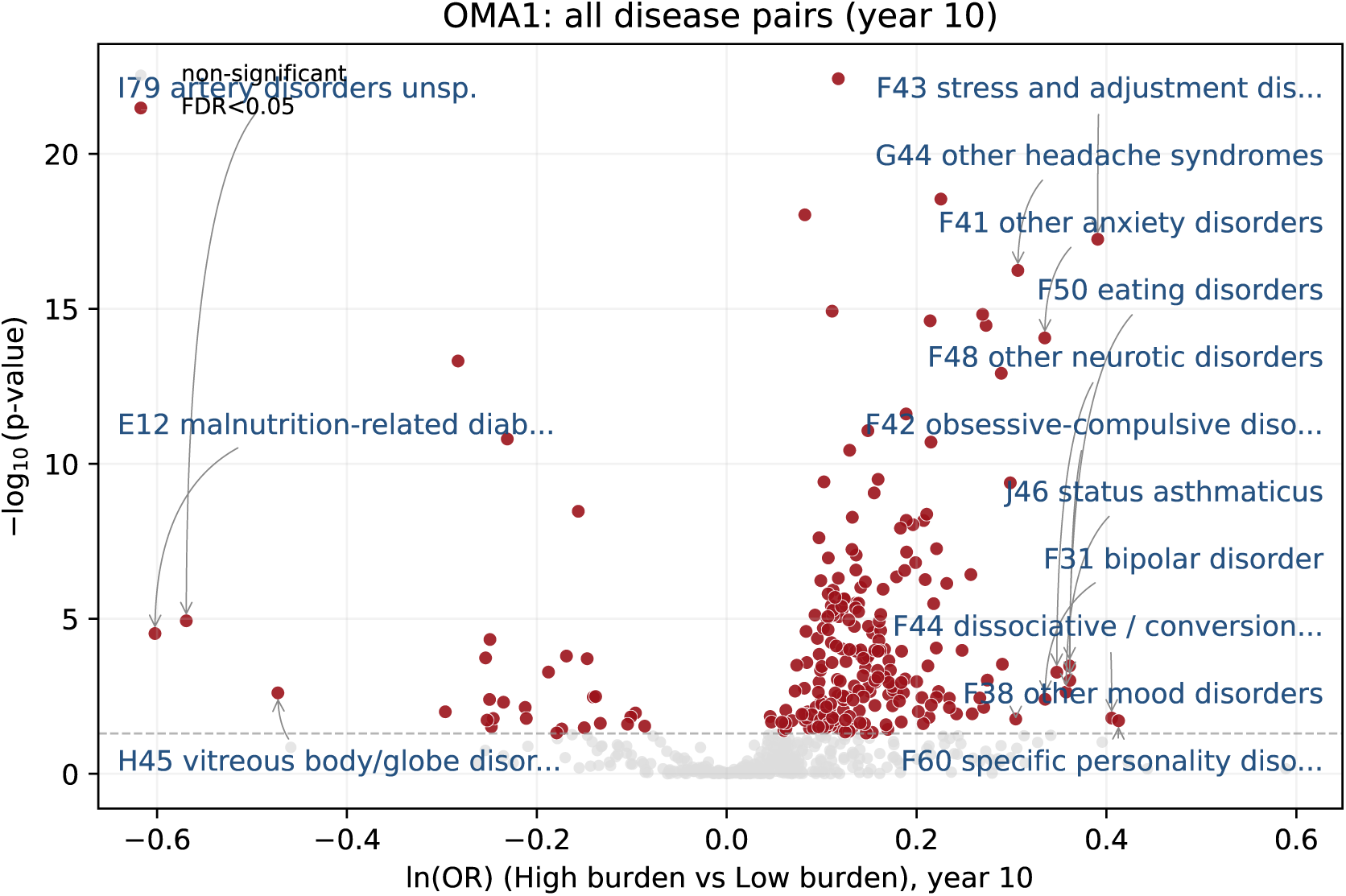
| OMA1 LoF burden associations across diseases at year 10. Scatter of ln(OR) versus. -log_10_(*pval*) at year 10 for all gene–disease pairs. Red: BH-FDR < 0.05; gray: non-significant. Labeled pairs satisfy BH-FDR < 0.05 and |ln(OR)| > 0.3 to keep visual clarity. ln(OR) compares predicted risk in high-burden vs low-burden (DeepRVAT score threshold of 0.75); significance is calculated with delta-method z-test on individual predicted risks between two groups with BH-FDR correction.

**Figure S13.**
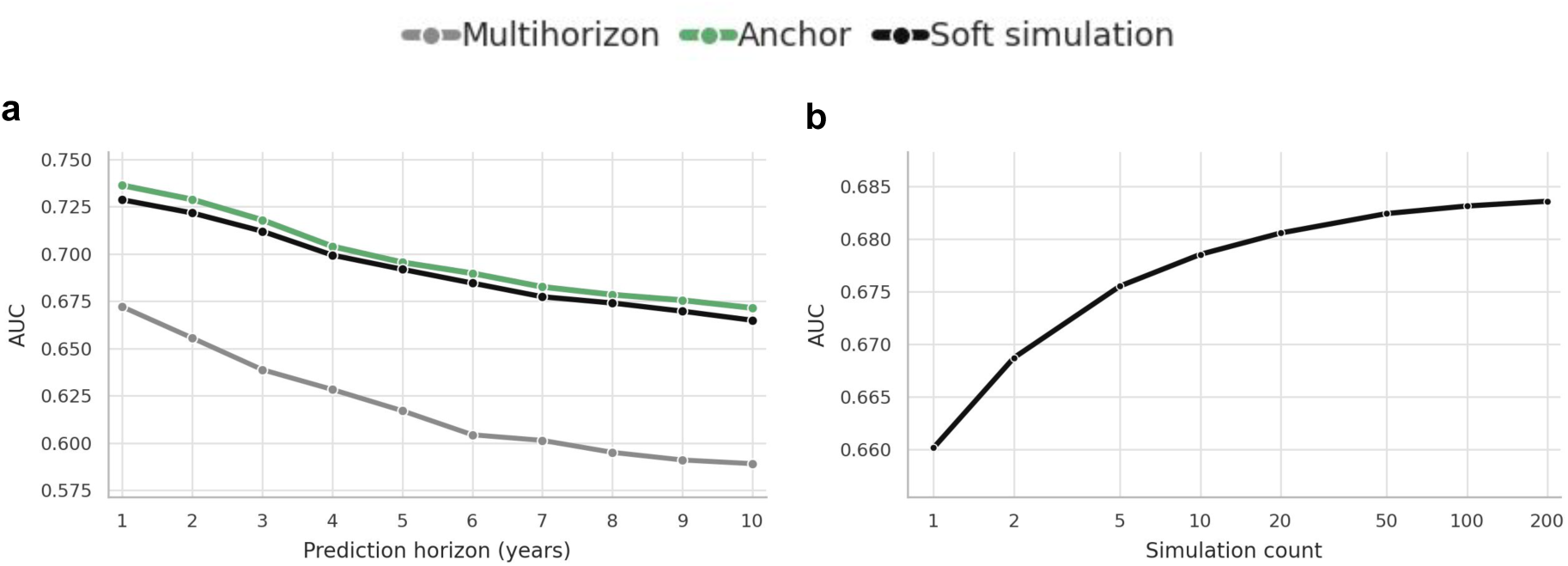
| Ablation of Delphi evaluation strategies and simulation convergence. (**a**) Absolute macro mean AUC across prediction horizons (1–10 years) for three evaluation schemes on the UK Biobank cohort: the *Multihorizon* estimator (standard offset-based next-event evaluation using the full diagnosis history); the *Anchor* estimator (history restricted to the baseline assessment, scoring the single next-token logit for disease-free individuals); and the *Soft simulation* estimator (forward simulation integrating continuous disease-probability quantities across sampled trajectories). (**b**) Convergence of the soft trajectory estimator as the number of simulated trajectories per individual increases from *n*=1 to *n*=200.

## B Supplementary Tables

**Table 1.** | Genes identified in the analysis, their biological functions, evidence categories, and supporting disease-related evidence from the literature.

| Gene | Evidence category | Main function | Established disease-related evidence |
| --- | --- | --- | --- |
| *HBB*<sup>70</sup> | Monogenic disease | β-globin subunit of hemoglobin | β-thalassemia; sickle-cell disease |
| *GCK*<sup>71</sup> | Monogenic disease | Glucose sensor in pancreatic β-cells and liver | GCK-MODY; fasting hyperglycemia |
| *LDLR*<sup>72</sup> | Monogenic disease | LDL cholesterol uptake receptor | Familial hypercholesterolemia |
| *APOB*<sup>73</sup> | Monogenic disease | Main apolipoprotein of LDL particles | Familial defective APOB-100; familial hypercholesterolemia |
| *PCSK9*<sup>74,75</sup> | Monogenic disease; protective LoF | Promotes LDLR degradation | Familial hypercholesterolemia; reduced LDL and CAD risk with LoF |
| *ABCA1*<sup>76</sup> | Monogenic disease | Cholesterol and phospholipid efflux to apoA-I | Tangier disease; HDL deficiency |
| *LCAT*<sup>77</sup> | Monogenic disease | Esterifies cholesterol in HDL | LCAT deficiency; fish-eye disease; renal disease |
| *ANGPTL3*<sup>78</sup> | Monogenic disease; protective LoF | Regulates triglyceride and cholesterol metabolism | Familial combined hypolipidemia; reduced cardiovascular risk |
| *SLC22A12*<sup>40</sup> | Monogenic disease; protective LoF | URAT1 renal urate reabsorption transporter | Renal hypouricemia; reduced gout risk with LoF |
| *SLC34A3*<sup>79</sup> | Monogenic disease | Renal sodium-phosphate cotransporter NaPi-IIc | Hypophosphatemic rickets with hypercalciuria |
| *COL4A3*<sup>80</sup> | Monogenic disease | Type IV collagen $\alpha 3$ chain in basement membrane | Alport syndrome; thin basement membrane nephropathy |
| *COL4A4*<sup>80</sup> | Monogenic disease | Type IV collagen $\alpha 4$ chain in basement membrane | Alport syndrome; thin basement membrane nephropathy |
| *CASR*<sup>81</sup> | Monogenic disease | Calcium-sensing receptor regulating PTH and calcium homeostasis | Familial hypocalciuric hypercalcemia |
| *TSHR*<sup>82</sup> | Monogenic disease | Thyroid-stimulating hormone receptor | Congenital hypothyroidism; hereditary hyperthyroidism |
| *TMPRSS6*<sup>83</sup> | Monogenic disease | Matriptase-2 involved in hepcidin and iron regulation | Iron-refractory iron-deficiency anemia |
| *KLF1*<sup>84</sup> | Monogenic disease | Erythroid transcription factor | Congenital dyserythropoietic anemia type IV |
| *TET2*<sup>85</sup> | Somatic variation | DNA demethylation in hematopoietic stem cells | Clonal hematopoiesis; myeloid malignancy risk |
| *ASXL1*<sup>85</sup> | Somatic variation | Chromatin and epigenetic regulator | Clonal hematopoiesis; myeloid malignancies |
| *SF3B1*<sup>86</sup> | Somatic variation | RNA splicing factor | Myelodysplastic syndrome; ring-sideroblast MDS |
| *CHEK2*<sup>87</sup> | Hereditary predisposition | DNA-damage checkpoint kinase | Breast and prostate cancer predisposition |
| *MC4R*<sup>88</sup> | Monogenic disease | Melanocortin receptor regulating appetite | Monogenic early-onset obesity |
| *KCNQ2*<sup>89</sup> | Monogenic disease | Neuronal voltage-gated potassium channel | Neonatal epilepsy spectrum |
| *LRP5*<sup>44</sup> | Monogenic disease | Wnt co-receptor regulating bone homeostasis | Osteoporosis-pseudoglioma syndrome |
| *G6PC1*<sup>90</sup> | Monogenic disease | Glucose-6-phosphatase catalytic subunit | Glycogen storage disease type Ia |
| *IGFALS*<sup>91</sup> | Monogenic disease | Stabilizes the circulating IGF-I-IGFBP-3 complex | Acid-labile subunit deficiency; short stature |

**Table 1 continued.**
| Gene | Evidence category | Main function | Established disease-related evidence |
| --- | --- | --- | --- |
| *SLC45A2*<sup>92</sup> | Monogenic disease | Melanosomal transporter involved in melanin synthesis | Oculocutaneous albinism type 4 |
| *OCA2*<sup>93</sup> | Monogenic disease | Melanosomal protein involved in pigmentation | Oculocutaneous albinism type 2 |
| *CLDN10*<sup>94</sup> | Monogenic disease | Tight-junction claudin involved in renal tubular ion transport | HELIX syndrome; salt-losing tubulopathy |
| *OMA1*<sup>45–47</sup> | Functional evidence | Mitochondrial stress-activated protease regulating OPA1 processing and mitochondrial dynamics | Experimental evidence links OMA1-mediated OPA1 cleavage to mitochondrial fragmentation and cellular stress responses |
| *PDE3B*<sup>95</sup> | Functional evidence | cAMP phosphodiesterase involved in insulin signalling, lipolysis, and energy homeostasis | Experimental and physiological evidence supports roles in glucose and lipid metabolism, adipocyte function, and insulin secretion |
| *SLC22A2*<sup>96</sup> | Pharmacogenetic evidence | Renal organic cation transporter OCT2 involved in tubular drug and metabolite uptake | OCT2 mediates renal transport and clearance of metformin, with functional variants influencing metformin pharmacokinetics |
| *PAPPA*<sup>97</sup> | Functional evidence | Metalloproteinase that cleaves IGF-binding proteins and regulates local IGF bioavailability | PAPP-A-mediated IGFBP-4 cleavage increases local IGF signalling involved in growth, tissue remodelling, and metabolism |

*Abbreviations:* CAD, coronary artery disease; LoF, loss of function; MDS, myelodysplastic syndrome. Evidence categories describe previously reported relationships and do not imply that variants identified in the present analysis share the same molecular consequence or mode of inheritance.

**Table 2.**
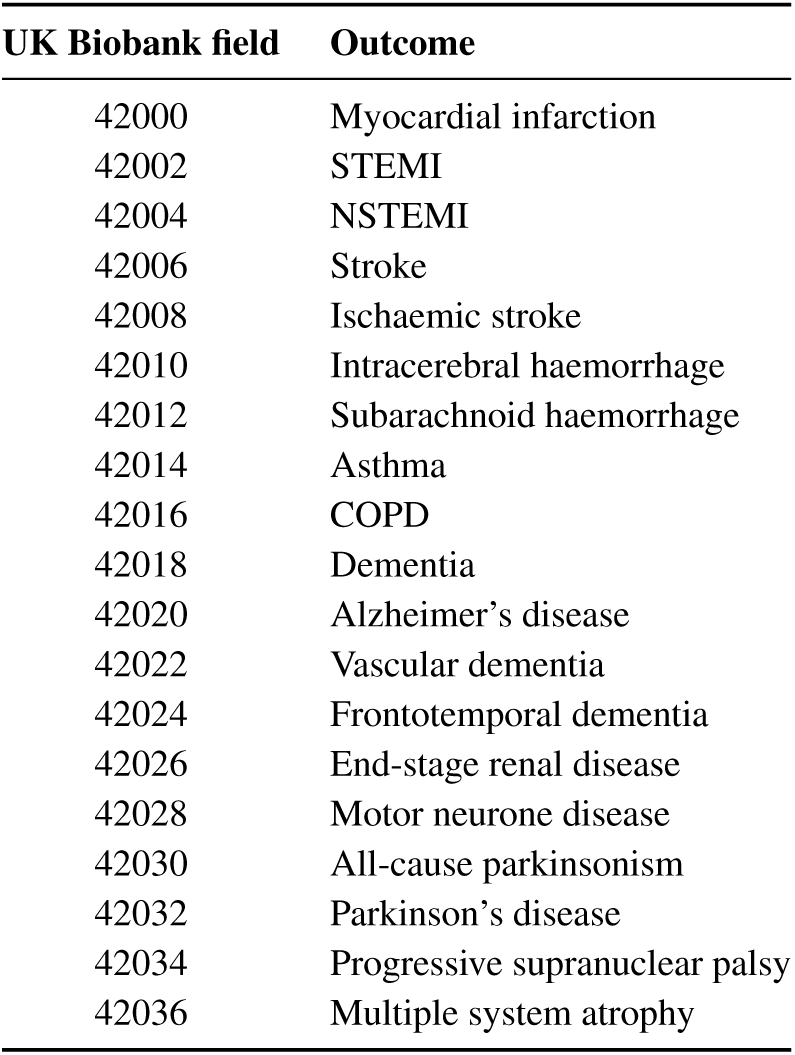
| Algorithmically defined outcomes from UK Biobank. UK Biobank field identifiers corresponding to algorithmically defined outcome variables. These outcomes were derived by UK Biobank through linkage to hospital admissions, death registrations, and other health records using predefined phenotyping algorithms.

**Table 3.** | Cohort characteristics of the UK Biobank cohort, by data split. Demographic summary of the study population, reported for the combined training and validation sets and separately for the held-out test set. Age corresponds to UK Biobank field 21003 (age when attended assessment centre). Sex (field 31) and self-reported ethnic background (field 21000) are shown as percentages. Ethnicity is aggregated to top-level UK Biobank categories; “Not reported” combines *prefer not to answer*, *do not know*, and missing responses.

| Characteristic | Train + Validation | Test |
| --- | --- | --- |
| *n* | 454,465 | 33,705 |
| **Age (years)** |  |  |
| Median | 58.0 | 58.0 |
| Range (min–max) | 37–73 | 39–70 |
| **Sex** |  |  |
| Female | 54.2% | 54.0% |
| Male | 45.8% | 46.0% |
| **Ethnic background** |  |  |
| White | 94.2% | 94.9% |
| Asian / Asian British | 1.9% | 2.1% |
| Black / Black British | 1.6% | 1.2% |
| Mixed | 0.6% | 0.5% |
| Chinese | 0.3% | 0.3% |
| Other ethnic group | 0.9% | 0.8% |
| Not reported | 0.5% | 0.4% |

**Table 4.** | Mapping of All of Us drinking frequency responses to UK Biobank field 1558 categories.

| All of Us response | UKB code |
| --- | --- |
| Prefer not to answer / Skip | –3 |
| 4 or more per week | 1 |
| 2 to 3 per week | 2 |
| 2 to 4 per month | 3 |
| Monthly or less | 5 |
| Never | 6 |

**Table 5.** | Mapping of All of Us smoking status responses to UK Biobank field 20116 categories.

| All of Us response | UKB code |
| --- | --- |
| Prefer not to answer / Skip / Don’t know | –3 |
| Not at all (currently) / Never smoked | 0 |
| Not at all, with a recorded quit age | 1 |
| Some days or every day | 2 |

**Table 6.** | Cohort characteristics of the All of Us cohort. Demographic summary of All of Us participants used for external validation. Age was computed from birth date and the index date (date of survey completion). Sex at birth and self-reported ehtnic background are shown as percentages. “Not reported” combines *None indicated* and *Skip* responses; *None of these* denotes participants who actively selected that no listed category applied to them.

| Characteristic | All of Us |
| --- | --- |
| *n* | 257,538 |
| **Age (years)** |  |
| Median | 55 |
| Range (min–max) | 25–81 |
| **Sex at birth** |  |
| Female | 68.0% |
| Male | 32.0% |
| **Ethnic background** |  |
| White | 56.83% |
| Black or African | 16.06% |
| Mixed | 4.38% |
| Asian | 2.30% |
| American Indian | 1.55% |
| Middle Eastern | 0.54% |
| Other | 1.05% |
| None of these | 1.05% |
| Not reported | 16.24% |

**Table 7.** | Hyperparameter search spaces and default values for XGBoost baseline models. Candidate values explored during Bayesian optimization of XGBoost baselines and fixed settings used for the default (non-optimized) baselines reported in the main text. Sweeps used W&B Bayes with 20 trials per (disease, horizon) for classification and 20 trials per disease for survival; the trial with highest validation AUC (classification) or Harrell’s *C*-index (survival) was retained. All models used tree_method=hist and up to 10,000 boosting rounds with early stopping after 20 rounds without validation improvement.

| Hyperparameter | Search space | Default value |
| --- | --- | --- |
| `max_depth` | {3, 5, 10} | 6 |
| `eta` | {0.01, 0.1, 0.3} | 0.3 |
| `min_child_weight` | {1, 5, 10} | 1 |
| `subsample` | {0.5, 0.75, 1} | 1 |
| `colsample_bytree` | {0.5, 0.75, 1} | 1 |
| `gamma` | {0, 1, 10} | 0 |
| `lambda` | {0, 1, 10} | 1 |
| `alpha` | {0, 1, 10} | 0 |

**Table 8.** | Matched disease set sizes for all statistical comparisons reported in the main text.

| Panel | Comparison | Cohort | *n* diseases |
| --- | --- | --- | --- |
| *Figure 2 — Performance across diseases and cohorts* |  |  |  |
| Fig. 2a | AUC 1y (RisQ vs. Delphi-2M, TabPFNv3, XGB × 4) | UKB | 327 |
|  | AUC 2y | UKB | 369 |
|  | AUC 5y | UKB | 419 |
|  | AUC 10y | UKB | 455 |
| Fig. 2b | C-index (RisQ vs. XGB Surv Opt/Default, Delphi-2M) | UKB | 486 |
| Fig. 2j | AUC 1y (RisQ vs. XGB Surv Opt, Delphi-2M) | AoU | 271 |
|  | AUC 2y | AoU | 271 |
|  | AUC 5y | AoU | 271 |
| Fig. 2k | C-index (RisQ vs. XGB Surv Opt, Delphi-2M) | UKB | 486 |
|  | C-index (RisQ vs. XGB Surv Opt, Delphi-2M) | AoU | 280 |
| Fig. 2* | AUC 2y (< 0.1% prevalence; RisQ vs. XGB Surv Opt) | UKB | 291 |
|  | AUC 10y (< 0.1% prevalence; RisQ vs. XGB Surv Opt) | UKB | 212 |
| *Figure 3 — Joint modeling across diseases, time, and modalities* |  |  |  |
| Fig. 3a | C-index, multi-disease vs. single-disease | UKB | 545 |
| Fig. 3b | C-index, multi- vs. single-disease, rarest 20% of diseases | UKB | 77 |
|  | rarest 40% | UKB | 193 |
|  | rarest 60% | UKB | 310 |
|  | rarest 80% | UKB | 428 |
|  | all diseases (100%) | UKB | 545 |
| Fig. 3c | C-index, uncapped (16y) vs. 2y supervision cap | UKB | 545 |
|  | uncapped vs. 5y supervision cap | UKB | 545 |
|  | uncapped vs. 10y supervision cap | UKB | 545 |
| Fig. 3e | Full model vs. single modality, per ICD-10 chapter (UKB) |  |  |
|  | II Neoplasms (C00–D49) | UKB | 93 |
|  | III Blood and immune (D50–D89) | UKB | 22 |
|  | IV Metabolic (E00–E89) | UKB | 34 |
|  | V Mental (F01–F99) | UKB | 48 |
|  | VI Nervous system (G00–G99) | UKB | 33 |
|  | VII Eye (H00–H59) | UKB | 22 |
|  | IX Circulatory (I00–I99) | UKB | 79 |
|  | X Respiratory (J00–J99) | UKB | 7 |
|  | XI Digestive (K00–K95) | UKB | 24 |
|  | XII Skin (L00–L99) | UKB | 52 |
|  | XIII Musculoskeletal (M00–M99) | UKB | 63 |
|  | XIV Genitourinary (N00–N99) | UKB | 66 |

**Table 9.** | Polygenic risk-score features included in the model. Standardized polygenic risk scores (PRSs) representing disease, anthropometric, biomarker, and reproductive phenotypes used as model input features. They are used to residualize predicted risks.

| Analysis variable | PRS phenotype |
| --- | --- |
| standard_prs_for_age_at_menopause_aam | Age at menopause (AAM) |
| standard_prs_for_age-related_macular_degeneration_amd | Age-related macular degeneration (AMD) |
| standard_prs_for_alzheimer's_disease_ad | Alzheimer's disease (AD) |
| standard_prs_for_asthma_ast | Asthma (AST) |
| standard_prs_for_atrial_fibrillation_af | Atrial fibrillation (AF) |
| standard_prs_for_bipolar_disorder_bd | Bipolar disorder (BD) |
| standard_prs_for_body_mass_index_bmi | Body mass index (BMI) |
| standard_prs_for_bowel_cancer_crc | Bowel cancer / colorectal cancer (CRC) |
| standard_prs_for_breast_cancer_bc | Breast cancer (BC) |
| standard_prs_for_cardiovascular_disease_cvd | Cardiovascular disease (CVD) |
| standard_prs_for_coeliac_disease_ced | Coeliac disease (CeD) |
| standard_prs_for_coronary_artery_disease_cad | Coronary artery disease (CAD) |
| standard_prs_for_crohn's_disease_cd | Crohn's disease (CD) |
| standard_prs_for_epithelial_ovarian_cancer_eoc | Epithelial ovarian cancer (EOC) |
| standard_prs_for_estimated_bone_mineral_density_t-score | Estimated bone mineral density T-score (eBMDT) |
| standard_prs_for_glycated_haemoglobin_hba1c_df | Glycated haemoglobin (HbA1c) |
| standard_prs_for_height_height | Height |
| standard_prs_for_high_density_lipoprotein_cholesterol_hdl | High-density lipoprotein cholesterol (HDL-C) |
| standard_prs_for_hypertension_ht | Hypertension (HT) |
| standard_prs_for_intraocular_pressure_iop | Intraocular pressure (IOP) |
| standard_prs_for_ischaemic_stroke_iss | Ischaemic stroke (ISS) |
| standard_prs_for_low_density_lipoprotein_cholesterol_ldl_sf | Low-density lipoprotein cholesterol (LDL-C) |
| standard_prs_for_melanoma_mel | Melanoma (MEL) |
| standard_prs_for_multiple_sclerosis_ms | Multiple sclerosis (MS) |
| standard_prs_for_osteoporosis_op | Osteoporosis (OP) |
| standard_prs_for_parkinson's_disease_pd | Parkinson's disease (PD) |
| standard_prs_for_primary_open_angle_glaucoma_poag | Primary open-angle glaucoma (POAG) |
| standard_prs_for_prostate_cancer_pc | Prostate cancer (PC) |
| standard_prs_for_psoriasis_pso | Psoriasis (PSO) |
| standard_prs_for_rheumatoid_arthritis_ra | Rheumatoid arthritis (RA) |
| standard_prs_for_schizophrenia_scz | Schizophrenia (SCZ) |
| standard_prs_for_systemic_lupus_erythematosus_sle | Systemic lupus erythematosus (SLE) |
| standard_prs_for_type_1_diabetes_t1d | Type 1 diabetes (T1D) |
| standard_prs_for_type_2_diabetes_t2d | Type 2 diabetes (T2D) |
| standard_prs_for_ulcerative_colitis_uc | Ulcerative colitis (UC) |
| standard_prs_for_venous_thromboembolic_disease_vte | Venous thromboembolic disease (VTE) |

## Supplementary Data

**Supplementary Dataset 1** - **List of diseases** Generated CSV file listing all prognosis targets used for RisQ disease-risk prediction and zero-shot evaluation: 1,246 outcomes comprising 588 clinician-curated targets, including ICD-10 hospital first-occurrence diagnoses, neoplasm chapter-prefix codes, and UK Biobank algorithmically defined outcomes, and 658 additional ICD-10 targets reserved for zero-shot evaluation. Columns: prognosis target index (id), UK Biobank field identifier where applicable (UKB field id), ICD-10 code where applicable (ICD code), disease or outcome name (name), and panel membership (target set: curated or additional).

**Supplementary Dataset 2** - **Female population cluster–disease enrichment results.** Generated CSV file list of the full filtered diseases enrichment across all clusters in the female population clustering (in the out-of-fold test set).

**Supplementary Dataset 3** - **Male population cluster–disease enrichment results.** This file contains the full filtered diseases and their enrichment in each cluster identified in the male population analysis (in the out-of-fold test set).

**Supplementary Dataset 4** –**Female age 50–55 cluster disease-risk enrichments.** Full results for disease-risk enrichment across clusters identified in the female age 50–55 England subcohort.

**Supplementary Dataset 5** – **Female age 50–55 cluster-associated features.** Full results for physiological, clinical, behavioral, and lifestyle features associated with clusters identified in the female age 50–55 England subcohort.

**Supplementary Dataset 6** – **Male age 50–55 cluster disease-risk enrichments.** Full results for disease-risk enrichment across clusters identified in the male age 50–55 England subcohort.

**Supplementary Dataset 7** – **Male age 50–55 cluster-associated features.** Full results for physiological, clinical, behavioral, and lifestyle features associated with clusters identified in the male age 50–55 England subcohort.

**Supplementary Dataset 8** - **Gene-level associations with the RisQ disease-risk representation.** This file lists the 32 genes significantly associated with the 40-PC RisQ disease-risk representation in the gene-level rare-variant burden analysis.

**Supplementary Dataset 9** - **Disease-specific associations for significant risk-structure genes.** This file contains disease-specific associations between DeepRVAT gene-level burden scores and 10-year predicted disease risks. The analysis included 32 genes and 588 diseases and identified 89 significant gene–disease pairs involving 29 genes; three genes yielded no disease-specific associations that met the reporting criteria. Statistical significance was assessed using a delta-method *z*-test comparing the log odds of the mean predicted disease risk between the high-burden and low-burden groups.

**Supplementary Dataset 10** - **List of features** Generated CSV file listing all 1,370 input features used to train RisQ at UK Biobank baseline assessment, grouped into nine training modalities. Columns: input feature index (id), UK Biobank field identifier (UKB field id), field description (name), training modality group (modality), and encoded value type (value type).

**Supplementary Dataset 11** - **List of medications** Generated CSV file listing all 2,505 GP prescription medications encoded as boolean input tokens in RisQ, derived from UK Biobank primary-care prescribing records and mapped to generic drug names with WHO ATC codes where available. Columns: medication token index (id), ATC code where applicable (ATC code), and canonical drug name (name).

